# Arterial Spin Labeling Reveals Persistent Cortical Hypoperfusion Linked to Cognitive Performance and Radiotherapy Dose in Post-Treatment Glioma Patients

**DOI:** 10.64898/2026.06.17.26354944

**Authors:** Joppe Van Rumst, Laurien De Roeck, Charlotte Sleurs, Sabine Deprez, Ahmed Radwan, Jan Petr, Kristien Bullens, Stefan Sunaert, Maarten Lambrecht

## Abstract

**Background:** Cognitive impairment is a prevalent long-term sequela in glioma patients, yet its cerebrovascular correlates remain poorly characterized. Arterial spin labeling (ASL) perfusion MRI offers a non-invasive means to quantify cerebral blood flow (CBF) and may serve as a sensitive correlate of radiotherapy (RT)-induced neurovascular injury.

**Methods:** Fifty WHO Grade 2/3 glioma patients and 50 matched healthy controls underwent pseudo-continuous ASL (pCASL) MRI and a standardized cognitive test battery. Regional CBF was compared between patients (n=44, after quality control) and controls (n=50) using ANCOVA with age, sex, and deep white matter CBF as covariates. In irradiated patients (~5 years post-RT), RT dose-CBF associations were assessed using region-wise regression. Additionally, regional CBF was compared among controls and low-dose (≤15 Gy) versus high-dose (≥40 Gy) regional RT exposure groups. Cognition-CBF associations were evaluated in a priori domain-specific regions of interest.

**Results:** Compared with controls, patients showed frontoparietal cortical hypoperfusion, with significantly lower CBF in middle frontal and superior/inferior parietal cortices (all *q*<0.01; *partial η*^*2*^=0.128-0.147). Region-wise regression showed no significant linear RT dose-CBF associations after correction. However, subgroup analyses identified RT dose-sensitive regions where the high-RT dose exposure group showed lower CBF than controls, most prominently in the left precentral and caudal middle frontal cortices (*q*<0.01; *adjusted-ΔCBF*≈-27.2--28.8 mL/100g/min). Perfusion in the left precentral and postcentral gyri of irradiated patients correlated positively with motor performance.

**Conclusions:** pCASL reveals persistent cortical hypoperfusion in glioma patients that spatially corresponds with RT dose exposure and associates with cognitive performance, positioning ASL as a promising non-invasive biomarker of RT-related neurovascular injury.

**Lay Summary:** Many patients treated for glioma experience long-term cognitive problems, but the underlying brain changes are not fully understood. In this study, we used a non-invasive brain imaging technique to measure blood flow in the brain years after treatment. We found reduced blood flow in specific brain regions, including areas that had received higher radiotherapy doses. Lower blood flow in motor brain regions was specifically linked to worse motor performance. These findings suggest that brain blood flow imaging may help detect treatment-related brain injury and better understand long-term side effects in glioma patients.

**Key Points:**

- Post-treatment glioma patients showed persistent frontoparietal hypoperfusion.
- High-RT dose exposure groups showed lower CBF than controls in select cortices.
- Higher left sensorimotor CBF was associated with better motor performance.

**Importance of the Study:** Cognitive decline following radiotherapy is a critical concern for glioma patients, who often face diagnosis in early adulthood and may live for decades with long-term treatment-related consequences. Despite growing interest in the vascular mechanisms of RT-induced neurotoxicity, in vivo cerebrovascular correlates of cognitive dysfunction in patients have been poorly characterized. This study demonstrates that non-invasive pCASL perfusion MRI detects persistent, spatially specific CBF alterations in clinically stable glioma patients that are linked to both RT dose distributions and cognitive performance. These findings position ASL as a clinically viable, contrast-free biomarker of RT-related neurovascular injury and provide a foundation for its integration into future longitudinal, multimodal studies and neuroprotective intervention trials aimed at preserving cognitive function in brain tumor patients.

## Introduction

Cognitive impairment is one of the most prevalent and disabling long-term sequelae in adult glioma patients, affecting more than half of patients who receive radiotherapy (RT) and spanning diverse cognitive domains including memory, executive functioning, attention, and motor performance.^1–3^ While the etiology of treatment-related neurocognitive decline is multifactorial, converging pre-clinical evidence implicates RT-induced cerebrovascular injury as a key mechanism, encompassing endothelial damage and microvascular occlusion.^4,5^ Despite growing mechanistic interest, the in vivo cerebrovascular correlates of cognitive dysfunction in glioma patients, and their spatial relationship to individualized RT dose distributions, remain poorly characterized.

Arterial spin labeling (ASL) perfusion MRI enables non-invasive, quantitative estimation of cerebral blood flow (CBF) by magnetically labeling water protons in arterial blood, without the need for exogenous contrast agents. Following standardization of pseudo-continuous ASL (pCASL) acquisition and post-processing,^6^ ASL has become an increasingly viable tool for detecting cerebrovascular alterations across a range of neurological conditions.^7^ In brain tumor patients, ASL has predominantly been applied in the acute or subacute treatment phase, where CBF reductions consistent with early vascular injury have been reported.^8–10^ Whether a distinct perfusion phenotype characterizes the chronic post-treatment phase, where late radiation effects, post-surgical remodeling, and hemodynamic compensation converge, and how such alterations relate to cognitive outcomes and RT dose, remains unknown.

Although evidence in glioma survivors is limited, studies in other oncological populations support a link between CBF and cognition. Lower or altered regional CBF has been associated with impairments in attention, processing speed, and memory.^11,12^ More specifically, longitudinal work in breast cancer patients has shown that treatment-related perfusion changes, particularly in frontal and sensorimotor cortices, correspond to cognitive difficulties shortly after chemotherapy.^13^ Similar patterns have been observed in a study of childhood non-CNS cancer survivors, in whom reduced perfusion in frontal, temporal, and hippocampal regions has been linked to poorer neurocognitive outcomes.^11^ Beyond oncology, longitudinal work in cerebral small vessel disease indicates that higher small vessel disease burden, together with associated brain hypoperfusion, predicts subsequent cognitive decline.^14^ Collectively, these findings demonstrate that CBF is a sensitive correlate of cognitive functioning across multiple disease contexts.

Cognitive impairment in long-term glioma survivors was previously associated with widespread structural and functional brain network disruption.^15–17^ These findings raise the question whether cerebrovascular dysregulation, measurable by ASL, independently contributes to cognitive dysfunction and spatially co-localizes with regions of high RT dose exposure in this survivor cohort. If so, CBF may represent a potential imaging biomarker of RT-induced neurotoxicity.

In the present study, we used pCASL MRI to: (1) characterize regional CBF differences between WHO Grade 2/3 glioma patients and individually age, sex and education matched healthy controls; (2) evaluate the relationship between regional CBF and local RT dose using region of interest (ROI)-wise regression, and assess regional CBF differences between controls and patients with low-RT dose (≤15 Gy) or high-RT dose (≥40 Gy) exposure; and (3) determine whether CBF in a priori domain-specific regions associates with performance across multiple cognitive domains. We hypothesized that patients would exhibit regionally aberrant CBF, that these abnormalities would show spatial correspondence with RT dose exposure, and that CBF in relevant neurocognitive networks would associate with cognitive performance.

## Materials and Methods

### Participants and Study Design

The study cohort comprised Dutch-speaking adult patients with WHO 2016 grade 2 or 3 gliomas, treated between 2007 and 2019 at the University Hospitals Leuven. To be eligible, patients had to be at least 1-year post-surgery and/or post-chemoradiotherapy, exhibit a WHO performance status of 0-2, and be in a stable phase of routine clinical follow-up. Key exclusion criteria were a history of other brain tumors, major psychiatric or neurological conditions, intellectual disability, or disease relapse following initial treatment. A group of healthy control participants was recruited through online forums and matched at the individual level based on sex, age (within a 3-year range), and educational level according to the Verhage classification system.^18^ A full overview of the population characteristics is available in *Supplementary Table S1*, which was adapted from our previous work.^15^

The study protocol was approved by the Research Ethics Committee of the University Hospitals Leuven (S63580), and all participants provided written informed consent in accordance with the Declaration of Helsinki. Data collection took place in a single session between 2021 and 2022, during which each subject underwent a comprehensive assessment including self-report questionnaires, standardized cognitive testing, and multimodal neuroimaging.

### Neuropsychological Assessment and Cognitive Domains

Cognitive functioning was evaluated across five domains using a standardized test battery. Language was assessed with the Controlled Oral Word Association Test,^19^ memory with the Hopkins Verbal Learning Test-Revised,^20^ and motor function with the Grooved Pegboard Test.^21^ Attention/processing speed and executive functioning were assessed using multiple instruments, including the Trail Making Test Parts A and B^22^, relevant WAIS-IV-NL subtests (Digit Span and Digit Symbol Substitution)^23^, and the Stroop Color and Word Test.^24^ The matrix reasoning subtest of the WAIS-IV-NL was used as a proxy measure of IQ (proxy IQ). Although not a direct measure of premorbid intelligence, it provides a reasonable estimate of general cognitive ability.

All cognitive test scores were converted to w-scores, adjusted for age and education based on the distribution of scores within the healthy control group. These w-scores were then averaged to derive composite scores for five cognitive domains and proxy IQ, with the grouping of tests into domains guided by established correlations and DSM-5 definitions (*Supplementary Table S2*). Full methodological details are described elsewhere.^15^

### MRI Data Acquisition

All MR imaging was performed on a 3T Philips Achieva scanner equipped with a 32-channel head coil. The structural protocol included a high-resolution T1-weighted magnetization-prepared rapid gradient echo (MPRAGE) sequence (0.8 mm isotropic voxel size, TR/TE 5800/2500 ms, flip angle 8°) and a 3D fluid-attenuated inversion recovery (FLAIR) sequence (1.0 mm isotropic voxel size, TR/TE 4800/340 ms). Perfusion imaging was performed using a 2D echo-planar imaging (EPI) pCASL sequence with 38 control-label pairs, a post-labeling delay of 1.8 s, a labeling duration of 1.8 s, and background suppression. The labeling plane was manually positioned perpendicular to the feeding arteries according to Alsop et al.^6^ Difference ASL images (ΔM = control-label) were generated on the scanner and extracted for further processing. A separate subject-specific M_0_ calibration image was not available for every participant. Detailed acquisition parameters are provided in *Supplementary Methods S1*.

### M_0_ Estimation and Calibration

Because subject-specific M_0_ maps were unavailable for all participants, we estimated a single arterial blood equilibrium magnetization constant (M_0,a_) using a separately acquired M_0_ calibration image obtained with the same readout but without background suppression.^25^ This calibration image was acquired from a high-quality adult reference subject who was not part of the study cohort. Cerebrospinal fluid (CSF) was used as the reference tissue,^26^ and CSF voxels were extracted from the M_0_ image using a FastSurfer (V7.3.2) derived CSF segmentation.^27^ Using literature values for the relaxation properties of CSF and arterial blood, we calculated M_0,a_ from the mean CSF signal.^25,28^ The full analytical derivation and validation is detailed in *Supplementary Methods S2*.

### Image Preprocessing

First, we performed semi-automated lesion segmentation of the resection cavity, tumoral tissue and/or perilesional gliosis, and ventricles using the T1-weighted and FLAIR images of the patients as previously described.^15^ A lesion distribution heatmap is visualized in *Supplementary Figure S2*.

All structural and ASL images were processed using ExploreASL *(V2*.*0*.*0 beta)*, a dedicated processing toolbox built on MATLAB (MathWorks, MA, USA, R2024b), Statistical Parametric Mapping 12 (SPM12, Wellcome Trust Centre for Neuroimaging, University College London, UK), and Diffeomorphic Anatomical Registration Through Exponentiated Lie algebra (DARTEL).^29^ Briefly, ΔM, T1-weighted, and FLAIR images were imported as sequence-sorted DICOMs using the import module, accounting for vendor-specific scale slopes, and subsequently structured in a brain imaging data structure (BIDS)-like ExploreASL format.

Structural T1-weighted images were segmented into gray matter (GM), white matter (WM), and CSF tissue probability maps using enhanced tissue priors. Spatial normalization to a population template in Montreal Neurological Institute (MNI) 152 standard space with 1.5 mm isotropic resolution was performed to support standardized visualization. For patient data, geodesic shooting with lesion cost-function masking was applied to prevent lesion voxels from influencing the registration process.^30,31^ To further avoid the influence of lesioned tissue on downstream analyses, GM and WM partial-volume probabilities were set to zero within lesion masks. This ensured that lesioned voxels did not contribute to ROI definitions, partial-volume correction, or regional CBF estimation.

CBF was quantified in native subject space from slice-time-corrected 2D-EPI pCASL data using a single-compartment model and calibrated with a universal M_0,a_ as detailed in the preceding section.^6^ Perfusion maps were coregistered to the structural image in native subject space and underwent partial-volume correction by linear regression using GM and WM tissue probability maps.^32^ Regional CBF values were sampled using the Desikan-Killiany-Tourville (DKT) atlas, as well as GM and deep WM (DeepWM) atlases transformed to native subject space.^33^ All ROI statistics are expressed as mean CBF within each region. The complete ExploreASL configuration file (DataPar.json) used for image processing is provided as *Supplementary File S1*.

For irradiated patients (n=33, *Supplementary Table S1*), three-dimensional RT dose maps were extracted from the clinical treatment planning system and registered to each subject’s native T1-weighted anatomy, as previously described.^17^ Regional mean RT dose was then sampled in native space using the same subject-space DKT atlas regions used for CBF extraction.

### Quality control and corrections

Completeness and quality checks were performed sequentially for all participants. From the 100 enrolled participants, comprising 50 patients and 50 healthy controls, two patients had incomplete ASL acquisitions due to scan session time constraints and were therefore excluded from perfusion analyses. Among the remaining participants with complete ASL data available, two additional patients were excluded after blinded visual inspection (J.V.R., S.S.) due to clear acquisition artifacts (e.g., poor signal-to-noise ratio, uneven labeling, motion, or arterial transit time (ATT)-related artifacts).

As an additional quantitative quality metric, we calculated the spatial coefficient of variation (sCoV = standard deviation (SD)/mean), a proxy for macrovascular and transit-related contamination, within GM.^29^ Using a conservative flag threshold of 0.60, we identified four patients with elevated sCoV (range 0.60-0.73) for targeted reinspection. Following reinspection, two patients were excluded due to vascular contamination (*sCoV* = 0.73) or motion artifacts (*sCoV* = 0.62), while the remaining cases showed moderate heterogeneity consistent with prior literature and were retained.^29^ This yielded a final perfusion analysis sample of 44 patients, including 33 who received RT, and 50 controls. Representative CBF maps and ΔM images of excluded participants, together with an example of an accepted CBF map, are shown in *Supplementary Figure S1*.

### Statistical analysis

All statistical analyses were performed using custom Python scripts (version ≥3.8), using standard libraries for data manipulation, statistical modeling, and visualization, including pandas, numpy, statsmodels, scipy, matplotlib, and seaborn. False discovery rate (FDR) correction was applied using the Benjamini-Hochberg method, with a significance threshold of q < 0.01 for the group-level CBF analysis and q < 0.05 for all other analyses. FDR-adjusted results are reported as q-values throughout the manuscript.

### Group-level CBF analysis

Group-level perfusion differences were examined at both global and regional levels using ANCOVA models. Whole-brain GM CBF (Total GM CBF) was compared between patients and controls with age, sex, and DeepWM CBF included as covariates, whereas DeepWM CBF was compared between patients and controls with age and sex included as covariates.

Regional CBF values, sampled from the DKT atlas, were compared between patients and controls using ANCOVA models including age, sex, and DeepWM CBF as covariates (*Supplementary Methods S3*). Partial η^2^ was reported as the effect size for the adjusted group effect. Regions were included if at least 20 participants in either group retained ≥1 mm^3^ of analyzable tissue (*Supplementary Table S6*). Model assumptions were evaluated using residual diagnostics (*Supplementary Section S4* and *Supplementary Figures S3-S4)*.

### Radiotherapy Dose-CBF Analysis

Using the regional mean RT dose values and paired regional CBF values from the same subject-space DKT regions, regional RT dose-CBF analyses were performed within the irradiated subgroup. To focus inference on regions with sufficient inter-individual RT dose variation, ROIs were included if the SD of regional mean RT dose across irradiated patients was ≥10 Gy (*Supplementary Table S8*).

RT dose-CBF associations were first modeled per ROI using ordinary least squares (OLS) regression in irradiated patients, predicting regional CBF from local mean RT dose while covarying for age, sex, and DeepWM CBF. This analysis tested whether regional CBF varied linearly with increasing local RT dose.

Because RT-related perfusion effects may be nonlinear or threshold-like, emerging primarily above clinically relevant RT dose levels, we additionally performed an ROI-wise three-group comparison. This analysis tested whether regional CBF differed between healthy controls and irradiated patients whose RT dose to that same ROI fell within a low-RT dose range (≤15 Gy) or high-RT dose range (≥40 Gy). Importantly, low- and high-RT dose exposure groups were defined separately for each ROI, based on each patient’s mean RT dose in that specific region. Thus, a patient could contribute to the high-RT dose group for one ROI and to the low-RT dose group for another ROI, depending on the regional RT dose distribution. Patients with intermediate regional RT doses (>15 and <40 Gy) were not included in the low-versus high-RT dose grouping for that ROI. For each eligible ROI, CBF was compared among healthy controls, the low-RT dose regional exposure group, and the high-RT dose regional exposure group using ANCOVA with age, sex, and DeepWM CBF as covariates. Adjusted group means were estimated for each ROI, and all pairwise contrasts were extracted. To avoid unstable estimates from sparse subgroup cells, formal inference was restricted to ROIs with at least eight subjects in each group. ROI-wise eligibility counts are provided in *Supplementary Table S10*.

As an additional exploratory sensitivity analysis, we examined whether regional DKT CBF was associated with clinical variables related to treatment burden and disease characteristics, including time since RT, tumor volume and chemotherapy exposure. These associations were assessed using partial Spearman correlations adjusted for age, sex, and DeepWM CBF with results summarized in *Supplementary Figure S6*. Full methodological details are provided in *Supplementary Methods S5*.

### Cognition-CBF Analysis

Associations between ROI-wise CBF and cognitive domain w-scores, including proxy IQ, were evaluated within the irradiated subgroup using partial Pearson correlations adjusted for DeepWM CBF. Multiple-comparison correction was applied within each cognitive domain. Analyses were restricted to a priori, domain-specific ROI sets obtained by mapping established neurocognitive networks to unilateral DKT regions (*Table 1*), with anatomical selection verified by radiologist A.R. For each domain, correlations were tested only within the corresponding a priori ROI set.

**Table 1.** A priori DKT ROIs per cognitive domain with canonical network assignments. **Caption**. A priori regions were selected to identify anatomically and functionally plausible unilateral DKT regions supporting the cognitive domains assessed in the present study, based on convergent evidence from functional, structural, and meta-analytic literature as cited. Domain labels reflect the study-specific cognitive constructs analyzed here and do not imply use of identical neuropsychological instruments across the cited source literature. The table is intended to justify hypothesis-driven ROI selection for cognition-CBF analyses rather than to define uniquely domain-specific brain regions. Reference numbers in the table correspond to the full citations listed in the references. DKT, Desikan-Killiany-Tourville; ROI, region of interest; L/R, left/right; dlPFC, dorsolateral prefrontal cortex; MD system, multiple-demand system; dACC, dorsal anterior cingulate cortex; ACC, anterior cingulate cortex; OFC, orbitofrontal cortex; SPL, superior parietal lobule; DAN, dorsal attention network; DMN, default mode network; RSC, retrosplenial cortex; MTL, medial temporal lobe; LEC/MEC, lateral/medial entorhinal cortex; mPFC, medial prefrontal cortex; IFG, inferior frontal gyrus; M1, primary motor cortex; S1, primary somatosensory cortex; SMA, supplementary motor area.

| Domain | DKT ROI (unilateral; L/R) | Canonical network / node | Sources |
| --- | --- | --- | --- |
| Proxy IQ | Caudal middle frontal L/R | dIPFC / MD system (fronto-parietal control) | 48,49 |
| Proxy IQ | Rostral middle frontal L/R | dIPFC / MD system (fronto-parietal control) | 48,49 |
| Proxy IQ | Caudal anterior cingulate L/R | dACC / cognitive control hub | 49,50 |
| Proxy IQ | Rostral anterior cingulate L/R | ACC / control | 49,50 |
| Proxy IQ | Insula L/R | Anterior insula / salience-control hub | 49,50 |
| Proxy IQ | Thalamus proper L/R | Thalamo-cortical relay (control loops) | 49 |
| Executive Function | Caudal middle frontal L/R | dIPFC / control | 49,50 |
| Executive Function | Rostral middle frontal L/R | dIPFC / control | 49 |
| Executive Function | Caudal anterior cingulate L/R | dACC / control | 49,50 |
| Executive Function | Rostral anterior cingulate L/R | ACC / control | 49 |
| Executive Function | Insula L/R | Anterior insula / salience | 49,50 |
| Executive Function | Lateral orbitofrontal L/R | Lateral OFC (context/valuation in control) | 49 |
| Executive Function | Medial orbitofrontal L/R | Medial OFC (valuation/context) | 49 |
| Executive Function | Thalamus proper L/R | Thalamo-cortical relay | 49 |
| Attention / Processing Speed | Superior parietal L/R | SPL / DAN parietal node | 51,52 |
| Attention / Processing Speed | Inferior parietal L/R | IPS/Angular / DAN-parietal interface | 51,52 |
| Attention / Processing Speed | Precuneus L/R | Precuneus / dorsal midline attention-DMN interface | 51 |
| Attention / Processing Speed | Caudal middle frontal L/R | FEF/dIPFC region / DAN frontal node | 51,52 |
| Attention / Processing Speed | Rostral middle frontal L/R | Dorsal frontal attention control | 51 |
| Attention / Processing Speed | Lateral occipital L/R | Occipital cortex / feature-based selection support | 51 |
| Motor Performance | Precentral L/R | Primary motor cortex (M1) | 41 |
| Motor Performance | Postcentral L/R | Primary somatosensory cortex (S1) | 42,53 |
| Motor Performance | Paracentral L/R | Medial motor (leg/foot; SMA proximity) | 54 |
| Memory | Hippocampus L/R | MTL episodic core (hippocampus) | 55 |
| Memory | Parahippocampal L/R | MTL (context/scene) pathway | 55 |
| Memory | Entorhinal L/R | MTL gateway (LEC/MEC) | 55 |
| Memory | Posterior cingulate L/R | Posterior DMN / memory retrieval hub | 56–58 |
| Memory | Isthmus cingulate L/R | RSC / posterior midline memory subsystem | 57 |
| Memory | Thalamus proper L/R | Hippocampal-thalamic interactions | 56 |
| Memory | Medial orbitofrontal L/R | mPFC/valuation node interfacing with DMN-memory | 58 |
| Language | Pars opercularis L/R | Inferior frontal (Broca's complex) | 59,60 |
| Language | Pars triangularis L/R | Inferior frontal (Broca's complex) | 59,60 |
| Language | Pars orbitalis L/R | Inferior frontal (IFG orbital; semantics/control) | 60 |
| Language | Superior temporal L/R | Posterior temporal (Wernicke's territory) | 59 |
| Language | Middle temporal L/R | Temporal semantic/lexical network | 60 |
| Language | Inferior temporal L/R | Temporal ventral stream (lexico-semantic) | 60 |
| Language | Supramarginal L/R | Inferior parietal (phonological buffer; dorsal stream) | 59,60 |

## Results

### Group-level CBF analysis

Globally, Total GM CBF was lower in patients than in controls (90.9 ± 20.1 vs. 98.6 ± 18.5 mL/100g/min, mean ± SD), with a significant group effect (*p* = 0.006, *partial η*^*2*^ = 0.082; *Supplementary Table S5*; diagnostic plots in *Supplementary Figure S5*). In contrast, DeepWM CBF itself did not differ between groups (*p* = 0.713, *partial η*^*2*^ = 0.002). Regionally, patients showed a frontoparietal pattern of lower CBF compared with controls (*Figures 1–2*). Significant group differences were observed in the left caudal middle frontal, bilateral rostral middle frontal, bilateral superior parietal, and left inferior parietal cortices (all *q =* 0.007; *partial η*^*2*^ = 0.128-0.147). The largest adjusted effect was observed in the left rostral middle frontal cortex (*partial η*^*2*^ = 0.147), with comparable effects across the remaining significant regions. Full ROI-level results are provided in *Supplementary Table S7*, with representative model diagnostics shown in *Supplementary Figures S3-S4*.

**Figure 1.**
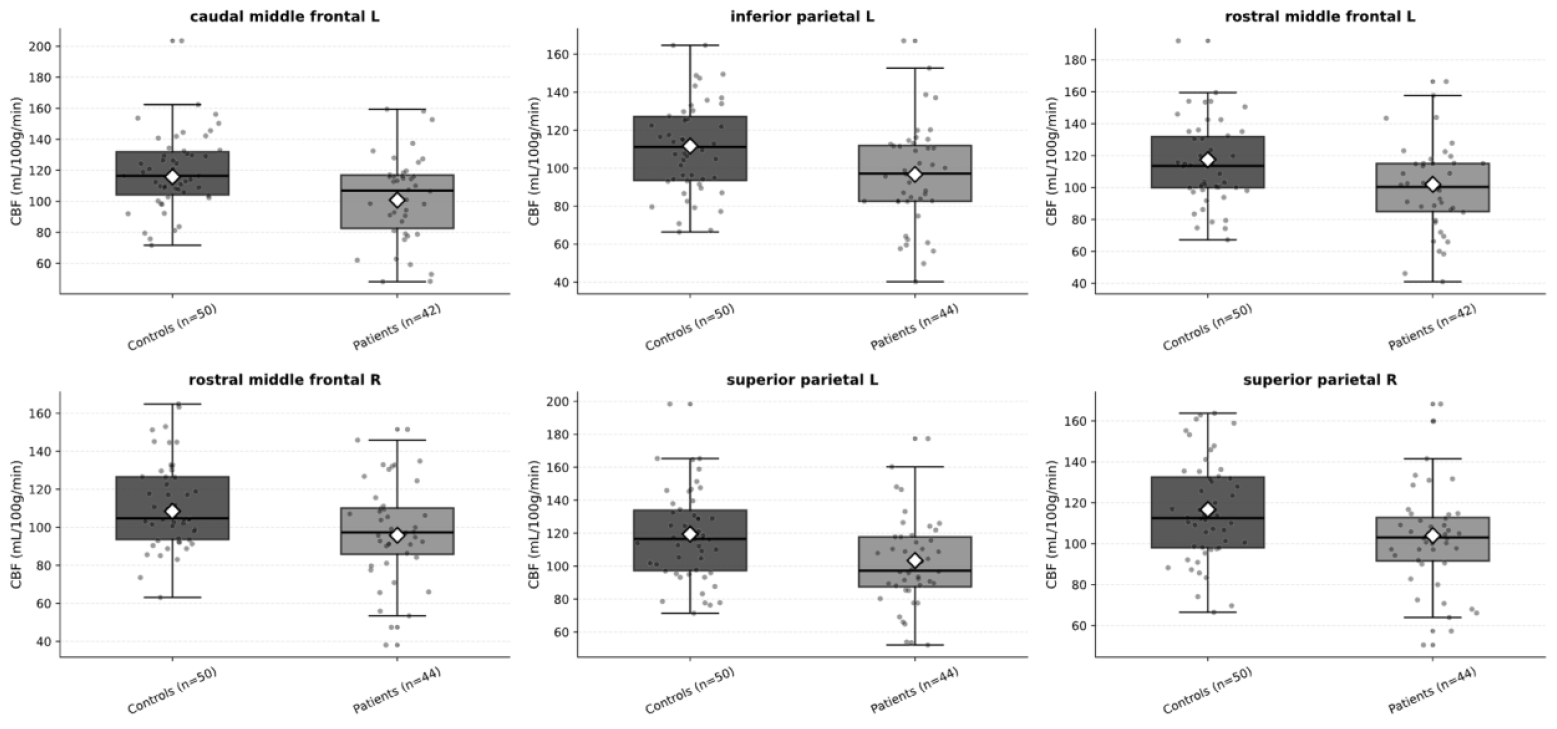
Combined boxplots and scatterplots of FDR-significant DKT regional CBF in patients (dark gray) versus controls (light gray). Each panel displays the distribution of cerebral blood flow (CBF, mL/100g/min) for a DKT region with ANCOVA FDR significance q < 0.01 (patients vs controls). Panels were ranked alphabetically on ROI name. Boxplots show group medians and interquartile ranges, with individual subject values overlaid as dots. White diamond indicates model-adjusted mean per group, estimated from ANCOVA (group: survivor vs control, covariates: Age, Sex, DeepWM CBF). X-axis labels denote group and sample size (N) per region after removing subject with insufficient ROI size (≤ 1 mm^3^). Region names follow DKT unilateral nomenclature (L/R). CBF, cerebral blood flow; DKT, Desikan-Killiany-Tourville; FDR, false discovery rate; q, Benjamini-Hochberg adjusted p-value.

**Figure 2.**
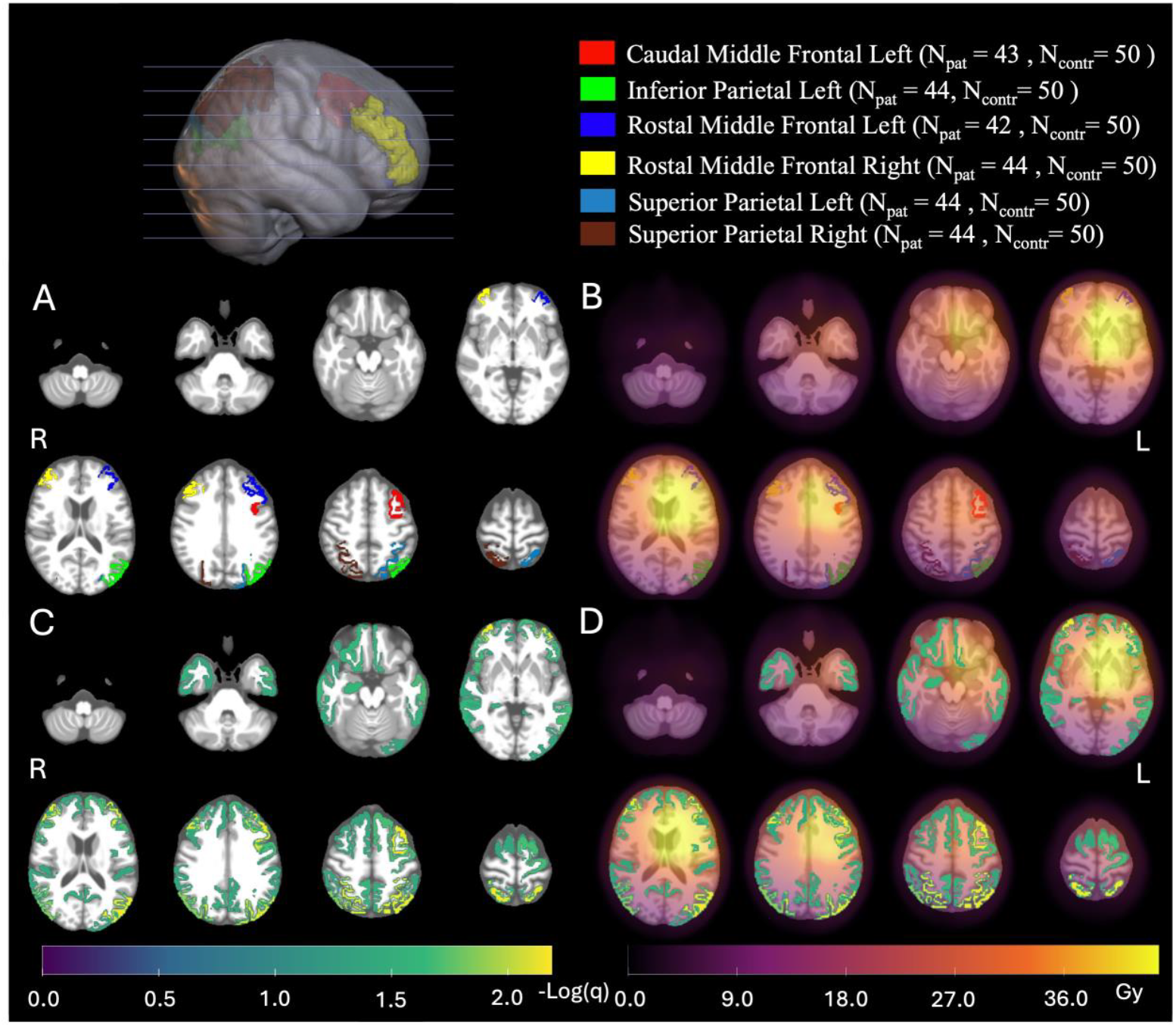
Significant DKT-region CBF differences and RT dose context in patients versus controls. (A) Binary overlays of DKT regions with ANCOVA FDR significance q < 0.01 (patients vs controls), displayed on the skull-stripped IXI555 MNI152 template. (B) The same regions as in (A) overlaid with the mean RT (Gy) for irradiated patients (n=33), registered to MNI space. (C) −log_10_(q) value map for all regions with q < 0.05. (D) The −log_10_(q) map from (C) overlaid with the mean RT dose map (Gy). Color bars beneath the figure denote −log_10_(q) (left) and mean RT dose (Gy) (right). A color-coded legend above the figure lists the significant DKT regions from panel (A) together with sample sizes per region (N_pat_, N_contr_) used after exclusions/quality control. Significance thresholds: q < 0.01 for panels A-B (primary) and q < 0.05 for panels C-D (secondary). q denotes FDR-adjusted p (Benjamini-Hochberg) from region-wise ANCOVA (group: survivor vs control; covariates: Age, Sex, DeepWM CBF). ROI labels follow DKT unilateral nomenclature (L/R). CBF, cerebral blood flow (mL/100g/min); DKT, Desikan-Killiany-Tourville; FDR, false discovery rate; MNI, Montreal Neurological Institute; N_pat_, number of patients; N_contr_, number of controls; RT, radiotherapy.

### Radiotherapy Dose-CBF Analysis

RT dose-CBF analyses showed a small number of nominal ROI-level associations in the OLS regression models, with the strongest uncorrected effects observed in the right thalamus proper (*slope* = 0.68 CBF/Gy, *p* = 0.002), right superior temporal cortex (*slope* = 0.50 CBF/Gy, *p* = 0.008), and left precentral gyrus (*slope* = −0.50 CBF/Gy, *p* = 0.010). These uncorrected effects were heterogeneous in direction, and none survived FDR correction (*Supplementary Table S9*).

In contrast, the ROI-wise three-group analyses revealed significant RT dose-sensitive differences in CBF. Significant contrasts between controls and high-RT dose regional exposure groups were observed in the left precentral gyrus (*q* < 0.001, *adjusted-ΔCBF* ≈ −27.2 mL/100g/min), left caudal middle frontal cortex (*q* = 0.001, *adjusted-ΔCBF* ≈ −28.8 mL/100g/min), right rostral middle frontal cortex (*q* = 0.016, *adjusted-ΔCBF* ≈ −19.1 mL/100g/min), and the left postcentral gyrus (*q* = 0.044, *adjusted-ΔCBF* ≈ −17.3 mL/100g/min). In the left precentral gyrus, CBF was also lower in the high-RT dose regional exposure group than in the low-RT dose regional exposure group (*q* = 0.021, *adjusted-ΔCBF* ≈ −23.8 mL/100g/min). In addition, the low-RT dose regional exposure group showed lower CBF than controls in the left inferior temporal cortex (*q* = 0.048, *adjusted-ΔCBF* ≈ −11.2 mL/100g/min). These group-level distributions and adjusted means are visualized in *Figure 3*. ROI-wise eligibility counts are summarized in *Supplementary Table S10*, and full adjusted means and pairwise contrast results are reported in *Supplementary Table S11*.

**Figure 3.**
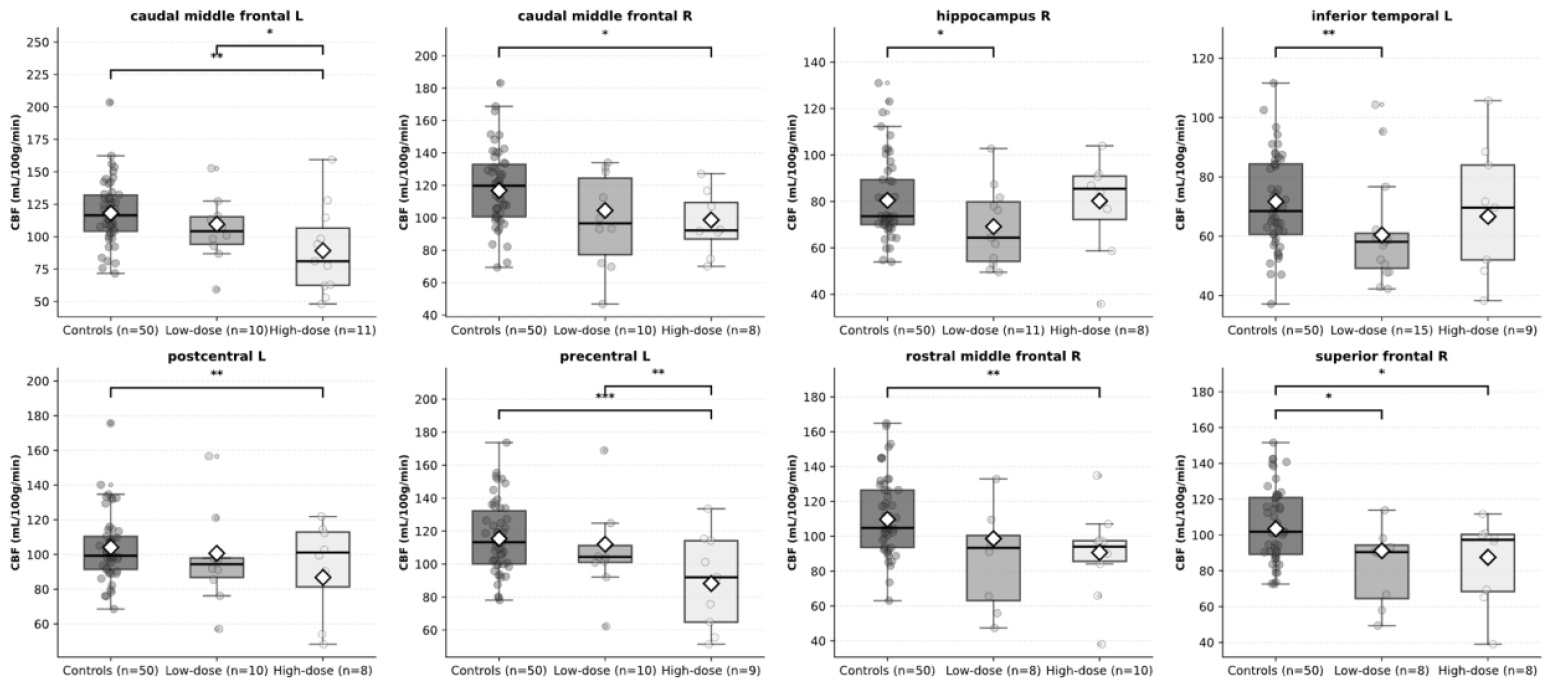
Combined boxplots and scatterplots of regional DKT CBF in ROIs with significant pairwise differences among controls and ROI-specific low- and high-RT dose exposure groups. Each panel displays the distribution of CBF (mL/100g/min) for a DKT region showing a significant pairwise contrast between RT dose regional exposure groups (ANCOVA, FDR-corrected q < 0.05). Panels include comparisons between controls (dark gray), low-RT dose (≤ 15 Gy, medium gray), and high-RT dose (≥ 40 Gy, light gray) groups as defined on an ROI-specific basis. Boxplots show group medians and interquartile ranges, with individual subject values overlaid as dots. White diamonds indicate model-adjusted mean CBF per group, estimated from ANCOVA models including Age, Sex, and DeepWM CBF as covariates. X-axis labels denote group and sample size (N) per region after exclusion of subjects with insufficient ROI volume (≤ 1 mm^3^). Panels are ordered alphabetically by ROI name. Region labels follow unilateral DKT nomenclature (L/R). Significance markers: * indicates p < 0.05 (uncorrected); ** indicates FDR significance (q < 0.05); *** indicates FDR significance (q < 0.01). CBF, cerebral blood flow; DKT, Desikan-Killiany-Tourville; FDR, false discovery rate; q, Benjamini-Hochberg adjusted p-value.

The exploratory clinical-variable sensitivity analysis identified a small number of uncorrected partial Spearman associations between regional perfusion and broader treatment- or disease-related variables. The strongest uncorrected effects were observed for chemotherapy exposure with the left lateral orbitofrontal cortex (*partial ρ* = −0.412, *p* = 0.026), and for tumor volume with the right parahippocampal cortex (*partial ρ* = 0.387, *p* = 0.034) and right pars triangularis (*partial ρ* = −0.380, *p* = 0.046). However, none of these associations survived FDR correction after adjustment for age, sex, and DeepWM CBF. These results are summarized in *Supplementary Figure S6 and Supplementary Table S12*.

### Cognition-CBF Analysis

The strongest CBF-cognition associations involved motor performance, which showed FDR-significant positive correlations with CBF in the left precentral gyrus (*r* = 0.69, *q* < 0.001) and left postcentral gyrus (*r* = 0.65, *q* < 0.001). Proxy IQ showed positive associations with CBF in the left caudal middle frontal, right caudal middle frontal, and right rostral middle frontal cortices (*r* = 0.435-0.441, *p* = 0.010 to 0.014), although these did not survive within-domain FDR correction (*q* = 0.058 for all three). Additional nominal positive associations were observed for executive function in the left caudal middle frontal and left caudal anterior cingulate cortex, for attention and processing speed in the left caudal middle frontal cortex, and for language in the left superior temporal cortex, left supramarginal gyrus, and left pars triangularis. These ROIs are visualized in *Figure 4* and reported in full in *Supplementary Table S13*.

**Figure 4.**
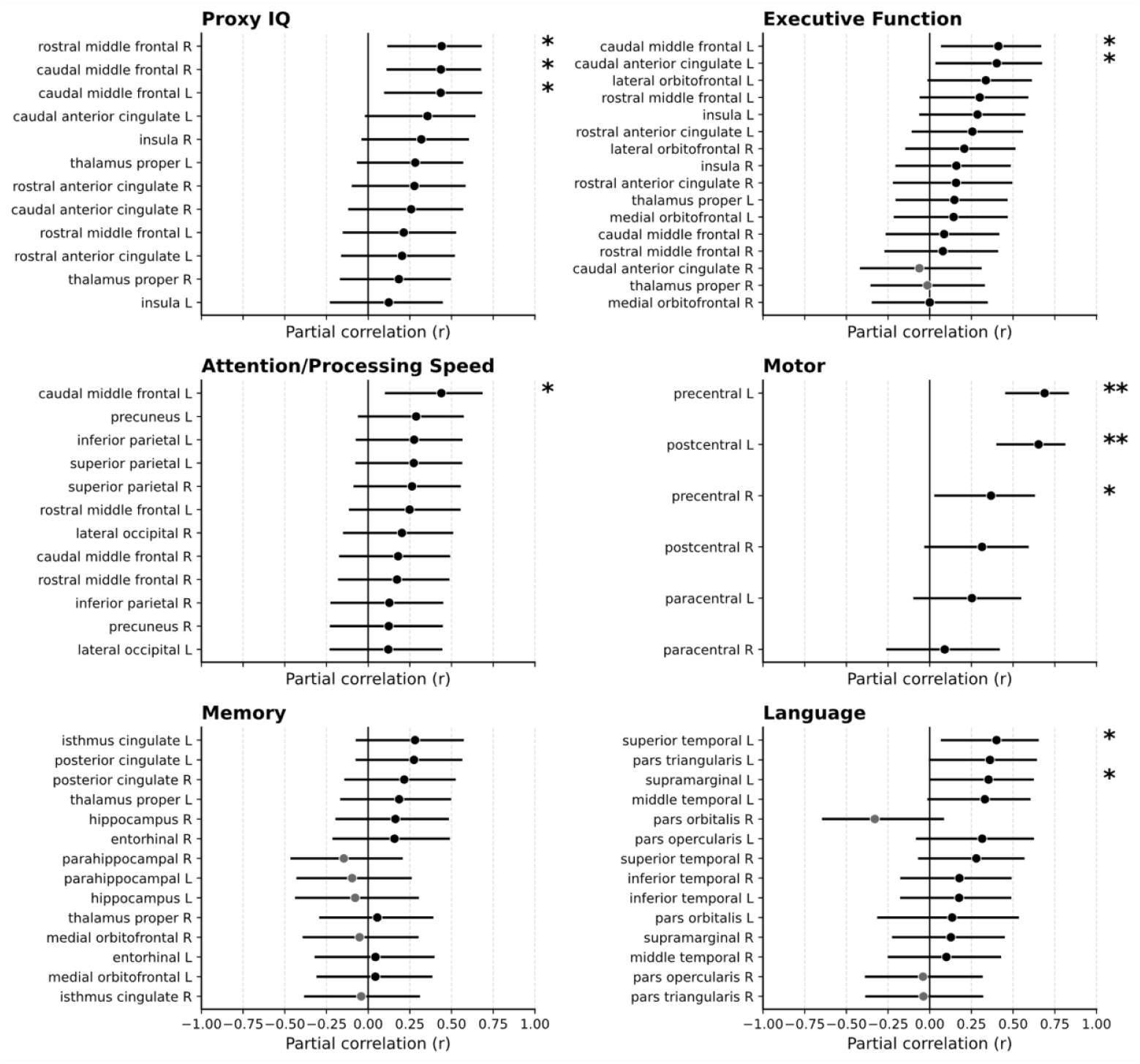
A priori ROI CBF-cognition associations in irradiated patients after adjustment for DeepWM CBF. Forest plots are arranged in a 3×2 grid (two columns × three rows), one panel per cognitive domain and Proxy IQ (Proxy IQ, Executive Function, Attention/Processing Speed, Motor, Memory, Language). Within each panel, points denote the partial Pearson correlation (r) between ROI-wise CBF and the domain w-score, with 95% confidence intervals (CIs) obtained via Fisher z-transformation; ROIs are ordered by absolute r. Intensity indicates direction of association (black = positive; gray = negative). Significance markers: * indicates p < 0.05 (uncorrected); ** indicates within-domain FDR significance (q < 0.05). ROI labels follow DKT unilateral nomenclature (L/R). CBF, cerebral blood flow (mL/100g/min); CI, confidence interval; DKT, Desikan-Killiany-Tourville; FDR, false discovery rate; RT, radiotherapy; q, Benjamini-Hochberg adjusted p-value.

## Discussion

To our knowledge, this is the first study to systematically characterize regional CBF using pCASL in clinically stable post-treatment WHO grade 2/3 glioma patients and to relate these perfusion alterations to regional RT exposure and multidomain cognitive performance. Patients showed a pattern of cortical hypoperfusion relative to controls. Irradiated patients were on average 5 years after RT (range 1.9 - 16.0 years), extending prior imaging studies reporting acute CBF reductions after cranial irradiation into the chronic post-treatment phase.^5,8^ This suggests that RT-related cerebrovascular alterations may represent an enduring, rather than transient, feature of the post-treatment brain in glioma patients.

Notably, the spatial distribution of hypoperfusion, centered on the middle frontal and parietal cortices, overlaps with regions previously identified in this same cohort as structural hubs on diffusion MRI-based structural connectivity analysis.^15^ While both sets of findings were obtained in the same sample, their convergence suggests persistent perfusion abnormalities and structural network disruption co-exist in topologically important brain regions. This co-localization is consistent with the idea that glioma treatment-related vascular injury, neuroinflammation, and white-matter damage may represent interrelated components of brain injury.^34^ Whether hypoperfusion precedes structural disconnection or develops in parallel as a distinct consequence of radiation exposure cannot be determined from the present cross-sectional data and will require longitudinal multimodal imaging. Nevertheless, the spatial convergence of perfusion and structural findings in network hubs reinforces the hypothesis that highly connected cortical regions are disproportionately vulnerable to treatment-related injury, a pattern with direct implications for RT planning strategies aimed at preserving critical network nodes.^15^

The RT dose-CBF analyses revealed a dissociation between continuous RT dose-response models and categorical RT dose-group comparisons. One interpretation is that radiation effects on brain perfusion may follow a threshold-like or nonlinear RT dose-response relationship, rather than a strictly linear association across the full regional RT dose range. This interpretation is supported by Price and colleagues, who observed RT dose-related CBF reductions confined to regions receiving more than 32 Gy, and by Petr et al., who similarly reported significant CBF decreases in high-RT dose regions (above 50 Gy) with negligible changes in low-RT dose regions (below 10 Gy).^8,35^ The absence of significant linear RT dose-CBF relationships in the present study may therefore reflect limited sensitivity of linear models to threshold-like effects, although limited statistical power and heterogeneous ROI-specific RT dose distributions may also have contributed.

Consistent with this threshold hypothesis, the three-group analysis identified a discrete set of cortical regions in which high-RT dose regional exposure groups showed lower CBF than healthy controls, most prominently in the left precentral, left postcentral, left caudal middle frontal, and right rostral middle frontal cortices. In the same regions no significant difference was observed between low-RT dose regional exposure groups and controls. This pattern suggests that the observed perfusion deficit is at least partially related to local cumulative RT dose rather than reflecting a generalized post-treatment effect. This interpretation is further supported by the left precentral gyrus, where CBF was lower in the high-RT dose regional exposure group than in the low-RT dose regional exposure group. This regional selectivity is supported by prior brain tumor imaging studies that demonstrated that radiation effects on normal-appearing brain tissue are anatomically heterogeneous. Nagtegaal et al. identified several cortical regions susceptible to RT dose-dependent thinning, whereas Seibert et al. reported greater vulnerability of higher-order cognitive regions and relative sparing of primary sensory and motor cortices.^36,37^ Importantly, our observation of hypoperfusion in the precentral and postcentral gyri does not necessarily contradict these structural findings, as cortical thickness and CBF capture different aspects of tissue injury. This interpretation is supported by Petr et al., who demonstrated that radiochemotherapy-related CBF reductions can occur independently of concurrent gray matter volume loss.^38^ Moreover, our ASL data were corrected for partial volume effects, reducing the likelihood that the observed hypoperfusion simply reflects local cortical atrophy.

A difference between the low-RT dose regional exposure group and controls was also observed in the left inferior temporal cortex. This effect was not accompanied by a significant high-RT dose versus control difference, a continuous RT dose-CBF association, or associations with clinical variables. Notably, the left inferior temporal cortex has been identified as a functional network hub in glioma patients and showed significant RT dose-cognition associations.^16,17^ These results indicate that different imaging modalities might provide complementary information on the post-treatment glioma brain and RT dose sensitivity.

The cognition-CBF associations were anatomically specific and largely consistent with findings from healthy populations.^39,40^ The strongest and most robust associations involved motor performance, which showed FDR-significant positive correlations with CBF in the left precentral and left postcentral gyri. This finding aligns with the known functional role of these sensorimotor regions in motor control and suggests that preserved perfusion in these areas is relevant to motor function in irradiated glioma patients.^41,42^

Notably, these same left sensorimotor regions also showed lower CBF in high-RT dose regional exposure groups compared with controls in the RT dose-group analysis. For the left precentral gyrus, this pattern was further supported by a significant high-RT dose versus low-RT dose difference. Although low-RT dose regions did not differ significantly from controls in these sensorimotor ROIs, the overlap between high-RT dose-associated hypoperfusion and motor-related CBF associations suggests that RT-related perfusion vulnerability in sensorimotor cortex may be functionally relevant. These findings complement previous structural connectome results from De Roeck and colleagues, in which network topology was linked to proxy IQ and attention,^15^ by suggesting that regional perfusion may provide an additional, vascular perspective on treatment-related brain changes.

In contrast, cognition-CBF associations outside the motor domain were weaker and did not survive within-domain FDR correction. Nominal associations involving frontal regions for executive function, attention/processing speed, and proxy IQ, and left temporal and parietal regions for language, may suggest broader links between regional perfusion and cognitive functioning. However, these findings require confirmation in larger, preferably longitudinal cohorts.

Multiple strengths of the present study should be highlighted. This study combined non-invasive perfusion imaging with individualized RT dose information, multidomain cognitive testing, matched healthy controls, and extensive quality-control procedures in a rare clinically stable post-treatment glioma cohort. The analytical pipeline integrated lesion-aware preprocessing, partial-volume correction, native-space ROI sampling, and individualized RT dose mapping, providing a structured framework for studying regional perfusion alterations in lesioned post-treatment brains. Notably, we validated a stable approach for M_0_ estimation and for handling an ASL dataset with only ΔM images, enabling accurate CBF quantification for regional and between-group analyses with wider applicability beyond the present cohort. Together with the use of a clinically feasible pCASL acquisition, the presented methods provide a rigorous and clinically meaningful framework for studying late treatment-related brain injury in glioma patients.

Several limitations should also be considered when interpreting the present findings. The cross-sectional design and absence of baseline pre-RT cognitive and perfusion data preclude conclusions about the causal contribution of treatment-related perfusion abnormalities to later cognitive outcome. The use of healthy volunteers rather than glioma patients treated without RT limits isolation of irradiation effects from surgery- and tumor-related influences. In addition, because subject-specific calibration images were unavailable, CBF quantification relied on a universal arterial M_0_ estimate derived from a single reference subject. Although this approach provides stable cohort-level scaling for the presented between-group and regional analyses, it does not correct for spatially varying acquisition factors such as receive-coil sensitivity and therefore does not support fully quantitative voxel-wise interpretation in individual subjects. A further limitation is the use of a single post-labeling delay ASL acquisition, which limits separation of true CBF from ATT effects. This is particularly relevant because arrival time of labeled blood may vary with both treatment-related vascular remodeling and age.^43^ This was partly mitigated through quality control and covariate adjustment, but future studies should consider multi-delay ASL to enable ATT-corrected CBF quantification and potentially improve sensitivity to RT-induced vascular alterations.^44^ Emerging ASL-based methods assessing blood-brain barrier water exchange may further clarify whether the observed abnormalities reflect altered perfusion, barrier dysfunction, or both.^45^ Finally, although the sample size was sufficient for patient-control and cognition-CBF analyses, it was more limited for regional RT dose-CBF analyses. Generalizability also warrants caution because 9 patients were diagnosed with isocitrate dehydrogenase 1 (IDH1) wild-type WHO 2016 grade 2 or 3 gliomas (mean follow-up of 2 years after RT), which would be classified as glioblastomas according to the current WHO 2021 criteria.

Future work should extend this vascular characterization using complementary MRI techniques such as susceptibility-weighted imaging (SWI) and quantitative susceptibility mapping (QSM), which can detect radiation-induced microbleeds, iron deposition, and changes in venous oxygenation as additional markers of microvascular injury.^46,47^ Whereas ASL provides a functional readout of perfusion, SWI and QSM offer structural and metabolic vascular information that may help clarify the biological basis of the CBF alterations observed here. Integrating these modalities with complementary diffusion-weighted imaging, resting-state functional MRI, individualized RT dose maps, and clinical variables would provide a more complete characterization of post-treatment brain tumors and could support the development of predictive models to identify patients at risk of later cognitive decline, with potential relevance for early intervention and more cognition-sparing treatment strategies.

## Conclusion

This study demonstrates that pCASL perfusion MRI reveals widespread cortical hypoperfusion in glioma patients, with regional patterns that partly correspond to RT dose exposure, and associated with cognitive performance. The spatial convergence of perfusion alterations with structural network hubs suggests that topologically central brain regions may be particularly vulnerable to treatment-related vascular injury. These findings position ASL perfusion MRI as a promising non-invasive biomarker of RT-related neurovascular injury, complementary to structural and functional connectivity measures, and support its inclusion in future longitudinal and multimodal studies of treatment-related cognitive decline in brain tumor patients.

## Data Availability

All data produced in the present study are available upon reasonable request to the authors

## Ethics Statement

The study was approved by the Research Ethics Committee UZ Leuven (S63580).

## Data Availability

The data supporting the findings of this study are available from the corresponding author upon reasonable request, subject to applicable ethical and institutional restrictions. Processing configuration files used for ASL analysis are provided as Supplementary Material.

## Author Contributions

J.V.R. analyzed the data, developed the analytical methods, and drafted the initial manuscript. L.D.R. conceived and designed the study and collected the data. S.S., A.R., and J.P. contributed to data curation and supervised the imaging analyses. M.L. secured funding for the study. M.L., S.D., L.D.R., C.S., and S.S. were involved in the conceptualization, formal analysis and supervision of the study. All authors (J.V.R., L.D.R., C.S., S.D., A.R., J.P., K.B., S.S., and M.L.) contributed to methodology design, interpretation of the findings, and critical revision of the manuscript, and approved the final version.

## Supplementary Materials

Supplementary_Information.pdf Supplementary_File_S1.json.zip

