## Supplementary Information for "Arterial Spin Labeling Reveals Persistent Cortical Hypoperfusion Linked to Cognitive Performance and Radiotherapy Dose in Post-Treatment Glioma Patients"

### Supplementary Materials

This document accompanies the manuscript " Arterial Spin Labeling Reveals Persistent Cortical Hypoperfusion Linked to Cognitive Performance and Radiotherapy Dose in Post-Treatment Glioma Patients".

#### Table of Contents

1. Supplementary Methods
    - S1. MRI Acquisition Parameters
    - S2. Detailed M0 Estimation and Sensitivity Analyses
        - S2.1. $T^*_2$ Sensitivity Analysis
    - S3. Deep White Matter CBF as a Global-Scaling Covariate in ASL Analysis
    - S4. Group-Level CBF Model Diagnostics
    - S5. Radiotherapy Dose-CBF Analysis
        - S5.1 Cohort and Imaging-Dosimetry Alignment
        - S5.2 ROI Eligibility Based on Regional RT Dose Variability
        - S5.3 Cross-Sectional (Between-Subject) Modeling
        - S5.4 ROI-Wise Three-Group Comparison
        - S5.5 Sensitivity Analysis
2. Supplementary Figures
    - Figure S1. Subject level examples used in quality control of ASL perfusion.
    - Figure S2. Lesion distribution heatmap
    - Figure S3. Diagnostic evaluation of regional CBF model in left caudal middle frontal.
    - Figure S4. Diagnostic evaluation of regional CBF model in left thalamus proper.
    - Figure S5. Group-level distributions of global perfusion indices.
    - Figure S6. Partial Spearman associations between regional DKT perfusion and clinical variables after adjustment for age, sex, and DeepWM CBF
3. Supplementary Tables
    - Table S1. Descriptive Characteristics of the Participants
    - Table S2. Neurocognitive tests used for cognitive domain composites and proxy IQ
    - Table S3. Summary of M0 Calibration Strategies and Outputs
    - Table S4. Full T2* Sensitivity Grid for Arterial M0
    - Table S5. Global Perfusion Comparison Between Patients and Controls
    - Table S6. DKT Regions Excluded Due to Insufficient Effective Volume
    - Table S7. DKT Regions Included in the Primary Perfusion Analysis
    - Table S8. ROI selection based on regional RT dose variability
    - Table S9. ROI-Wise OLS Regression of Dose-CBF Associations
    - Table S10. ROI-wise sample sizes and eligibility for the three-group RT dose comparison
    - Table S11. ROI-wise three-group ANCOVA of regional CBF by RT dose exposure group

Table S12. Partial Spearman correlations between regional DKT perfusion and clinical variables in irradiated patients
Table S13. A priori ROI cognition-CBF partial correlations in irradiated patients
4. Supplementary References

### 1. Supplementary Methods

#### S1. MRI Acquisition Parameters

All magnetic resonance imaging was performed on a 3T Philips Achieva scanner equipped with a 32-channel head coil. The structural protocol included a high-resolution T1-weighted magnetization-prepared rapid gradient echo (MPRAGE) acquisition (0.8 mm isotropic voxel size, repetition time/echo time (TR/TE) 5800/2500 ms, inversion time 1000 ms, flip angle 8°, field of view 320 × 320 mm) and a 3D fluid-attenuated inversion recovery (FLAIR) sequence (1.0 mm isotropic voxel size, TR/TE 4800/340 ms, inversion time 1650 ms, flip angle 90°, field of view 256 × 256 mm).

The primary perfusion sequence was a 2D echo-planar imaging (EPI) pseudo-continuous arterial spin labeling (pCASL) acquisition with 38 control-label pairs, a post-labeling delay of 1.8 s, a labeling duration of 1.8 s, and background suppression using two pulses (1.813/3.135 s). Imaging was performed with an acquired voxel size of 1.875 × 1.875 × 6.0 mm, TR 4.39 s, and TE 12.54 ms. The phase-encoding direction was anterior-to-posterior, with an effective echo spacing of 0.139 ms and a total readout time of 17.64 ms. Difference images ($\Delta M$ = control-label) were calculated on the scanner and extracted for further processing. A separate subject-specific M0 calibration image was not acquired for every subject.

The labeling plane was positioned using a multi-planar localizer to identify distal internal carotid and vertebral arteries. The plane was placed approximately 90 mm inferior to the center of the imaging volume and oriented perpendicular to the dominant flow direction, avoiding the carotid bifurcation, vessel curvature, and susceptibility-prone regions. Vendor visualization of the four feeding arteries was used to verify correct placement prior to acquisition.[1] Procedures were kept consistent across all participants.

#### S2. Detailed M0 Estimation and Sensitivity Analyses

Because the pCASL acquisition used background suppression and individual $M_0$ maps were not acquired for every study participant, we estimated a universal arterial blood equilibrium magnetization ($M_{0,a}$) from a high-quality reference subject. The first $M_0$ control volume was used, and cerebrospinal fluid (CSF) signal was sampled using three strategies:

(i) a high-probability CSF tissue map (pCSF ≥ 0.90),
(ii) a ventricular CSF mask derived from FastSurfer[2], and
(iii) their intersection (ventricular ∩ pCSF ≥ 0.90).

Quantification followed:

$$M_{0,a} = \frac{< M_{0,CSF} > e^{TE(\frac{1}{T^*_{2,CSF}} - \frac{1}{T^*_{2,a}})}}{\lambda_{CSF}}$$

Where $< M_{0,CSF} >$ denotes the mean $M_{0,CSF}$ signal intensity, TE is the echo time, $T^*_{2,CSF}$ and $T^*_{2,a}$ are the effective transverse relaxation times of CSF and arterial blood, respectively, and $\lambda_{CSF}$ is the CSF-blood partition coefficient.[3]

Using TE = 12.542 ms and literature-based values ($T^*_{2,CSF}$ = 400 ms; $T^*_{2,a}$ = 47 ms, $\lambda_{CSF}$ = 1.15 ml/g), the exponential correction factor was 0.790.[3,4] The resulting $M_{0,a}$ values for all three sampling strategies are shown in *Supplementary Table S3*. Based on visual inspection, the intersection strategy, defined by the overlap between the high-probability CSF map and a ventricular CSF mask, was selected to minimize partial-volume contamination. Using this approach, $M_{0,a}$ was estimated as 61.44 × $10^3$ a.u. This result was further used in the analysis.

To assess physiological plausibility, mean tissue values were also extracted from the same calibration volume. These were $M_{0,CSF}$ = 89.42 × $10^3$, $M_{0,GM}$ = 74.29 × $10^3$ and $M_{0,WM}$ = 68.59 × $10^3$, yielding the expected proton density ordering CSF > gray matter (GM) > white matter (WM) and thereby supporting the plausibility of the calibration target.[5]

##### S2.1. $T^*_2$ Sensitivity Analysis

Using the ventricular ∩ pCSF strategy, we evaluated physiologically plausible values of $T^*_{2,CSF} \in \{400, 450, 500, 550, 600, 650, 700\}$ ms and $T^*_{2,a} \in \{40, 45, 50, 55, 60\}$ ms. Across this grid, $M_{0,a}$ ranged from 57.86 × $10^3$ a.u. (700/40 ms) to 65.10 × $10^3$ a.u. (400/60 ms), relative to the reference estimate of 61.44 × $10^3$ a.u. which was used, corresponding to a variation of ±5.9%. The complete parameter grid is provided in *Supplementary Table S4*.

The use of a universal $M_0$ calibration standardizes ΔM to arterial equilibrium magnetization at the subject level, but it does not correct for spatially varying factors such as receive-coil sensitivity, local proton-density differences, or slice-profile variation. Accordingly, the derived CBF maps are most appropriate for group-level comparisons and regional statistical analyses, rather than for absolute voxelwise quantification within an individual subject.

#### S3. Deep-White-Matter CBF as a Global-Scaling Covariate in ASL Analysis

Deep white-matter (DeepWM) CBF was included as an empirical proxy for subject-level global ASL scaling, capturing three potential sources of inter-individual variability that the universal $M_{0,a}$ calibration does not fully address.[3] First, and most importantly, labeling efficiency (α) varies across individuals as a function of carotid geometry, blood flow velocity, and cardiac output; because α scales the magnetization difference signal (ΔM) globally and independently of $M_0$, it produces a proportional bias across all brain regions in a given subject.[3,6] Second, the fixed background suppression pulse timings interact with individual tissue T1 values, introducing residual signal variability in ΔM that likewise operates as a global multiplicative factor.[1] Third, interindividual differences in hematocrit alter both blood water proton density and blood T1, meaning the universal $M_{0,a}$ is precisely applicable only to subjects whose hematocrit matches the reference individual.[7]

To help mitigate these effects, we adopted a WM reference approach, a practice commonly used in dynamic susceptibility contrast MRI, where normal-appearing white matter serves as stable scaling factor because of its relatively low and consistent perfusion.[8,9] In this cohort DeepWM CBF showed limited absolute variability (standard deviation (*SD)*: controls 3.11, patients 3.71 mL/100g/min) and importantly, did not differ between patients and controls (ANCOVA adjusted for age and sex: $p = 0.713$) (*Supplementary Table S5*). These observations support the use of DeepWM CBF as a scaling covariate in the present analysis, while acknowledging that its reproducibility across different scanners and its moderate signal-to-noise ratio might affect implementation in other settings.

#### S4. Group-level CBF Model diagnostics

To assess whether the assumptions of the linear models were tenable, we visually inspected standard diagnostic plots for the global CBF model and selected representative regional models. Residual normality was evaluated using quantile-quantile plots and residual distributions. These diagnostics did not indicate major systematic deviations from approximate normality. Residuals plotted against fitted values did not show clear heteroscedasticity, curvature, or other obvious patterns suggestive of model misspecification. The linearity of continuous covariate effects was evaluated using diagnostic plots against age and DeepWM CBF, which did not suggest pronounced nonlinear trends in the inspected models. Leverage and standardized-residual plots were used to screen for influential observations, and no single observation appeared to exert disproportionate influence. Representative diagnostic panels are shown for one significant ROI, the left caudal middle frontal cortex, and one non-significant ROI, the left thalamus proper, in *Supplementary Figures S3 and S4*.

#### S5. Radiotherapy Dose-CBF Analysis

##### S5.1 Cohort and Imaging-Dosimetry Alignment

RT dose-CBF analyses were restricted to irradiated patients ($n = 33$) for whom individualized three-dimensional dose distributions were available from the clinical treatment planning system. For each patient, the planning CT and associated dose grid were exported and registered to the native T1-weighted anatomical image, as previously described.[10] Registration quality was visually inspected to ensure consistent alignment at tissue and ventricular boundaries.

Regional mean absorbed radiotherapy (RT) dose was extracted in native space for unilateral Desikan-Killiany-Tourville (DKT) atlas regions using the same subject-space masks employed for partial-volume-corrected CBF quantification. This yielded a matched matrix of regional mean RT dose and regional CBF for each subject and ROI.

##### S5.2 ROI Eligibility Based on Regional RT Dose Variability

To restrict analyses to regions that exhibited sufficient inter-individual RT dose variability, ROIs entered the RT dose-CBF analysis pipeline if their across-patient RT dose standard deviation was ≥ 10 Gy. This criterion ensured that RT dose-CBF associations were evaluated only in regions with adequate variability to support regression and group-based inference. The final set of eligible ROIs is listed in *Supplementary Table S8*.

##### S5.3 Cross-Sectional (Between-Subject) Modeling

Cross-sectional associations between RT dose and CBF were evaluated on an ROI-by-ROI basis using ordinary least squares (OLS) regression. For each eligible ROI $r$, regional CBF was modeled as a function of regional mean RT dose, with age, sex, and DeepWM CBF included as covariates:

$$\mathrm{CBF}_{i,r} = \beta_{0,r} + \beta_{1,r}\,\mathrm{Dose}_{i,r} + \beta_{2,r}\,\mathrm{Age}_i + \beta_{3,r}\,\mathrm{Sex}_i + \beta_{4,r}\,\mathrm{DeepWM}_i + \varepsilon_{i,r}.$$

The coefficient $\beta_{1,r}$ was taken as the estimate of the dose-CBF association for each ROI. Standard errors, t-statistics, and 95% confidence intervals were extracted. Significance across ROIs was controlled using Benjamini-Hochberg false discovery rate (FDR) correction applied at q = 0.05. Complete ROI-level outputs are provided in *Supplementary Table S9*.

##### S5.4 ROI-Wise Three-Group Comparison

Because radiation-related perfusion effects may be nonlinear or threshold-like, emerging primarily above clinically relevant RT dose levels, we additionally implemented an ROI-wise three-group comparison framework. For each ROI, irradiated patients were classified into low-RT dose (≤15 Gy) or high-RT dose (≥40 Gy) regional exposure groups based on the regional RT dose in that specific region and were compared with healthy controls.

Group differences in regional CBF were assessed using analysis of covariance (ANCOVA), with age, sex, and DeepWM CBF included as covariates. For each ROI, adjusted group means were estimated and all pairwise contrasts (control vs low-RT dose, control vs high-RT dose, and high-RT dose vs low-RT dose) were extracted. Statistical significance across ROIs was controlled using Benjamini-Hochberg FDR correction applied at q = 0.05.

To ensure stable estimation, only ROIs with at least eight subjects per group were considered for formal inference. The full ROI-wise eligibility counts are provided in *Supplementary Table S10* and ROI-wise adjusted means and pairwise contrast results are reported in *Supplementary Table S11*.

##### S5.5 Sensitivity Analysis

To explore whether CBF variation was related to treatment- and tumor-related clinical factors beyond regional RT dose, we performed an additional ROI-wise exploratory analysis within the irradiated subgroup. For each unilateral DKT region, partial Spearman correlations were computed between regional CBF and the following clinical variables: time since RT (years), tumor volume ($cm^3$), and chemotherapy exposure (coded 0/1). Partial correlations were adjusted for age, sex, and DeepWM CBF to account for demographic effects and subject-level global perfusion scaling. Only unilateral DKT regions with at least 10 available observations were included. Statistical significance was summarized using raw p-values and Benjamini-Hochberg FDR correction applied within each predictor across ROIs. The results are visualized in *Supplementary Figure S6* and *Supplementary Table S12*.

#### 2. Supplementary Figures

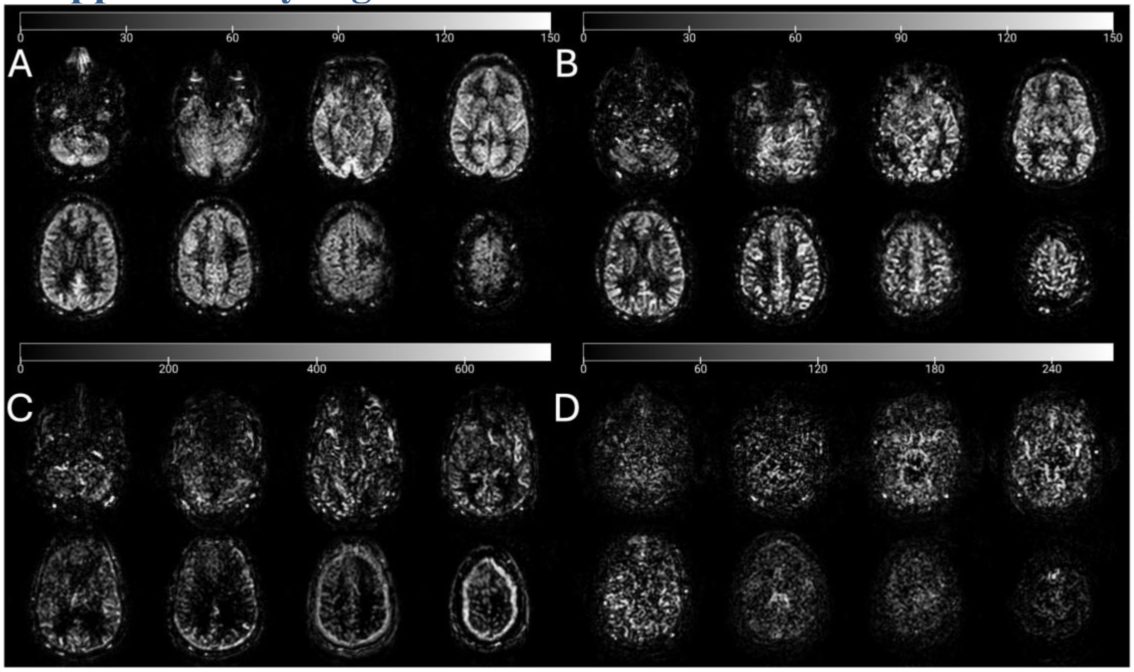

**Figure S1. Subject-level examples used in quality control of ASL perfusion.**
(A) Representative participant with acceptable CBF image (2D-EPI pCASL), used as reference for visual quality control; color bar shows CBF (mL/100g/min). (B) Participant excluded after targeted re-inspection for macrovascular/transit contamination (sCoV = SD/mean = 0.729); color bar shows CBF (mL/100g/min). Elevated sCoV on single-post-labeling-delay ASL is a recognized proxy for arterial-transit/intravascular signal contamination. (C-D) Two participants removed before analysis due to severe acquisition artifacts on the raw ΔM images (difference of control-label); color bars are dimensionless (ΔM units). All maps are shown in subject/native space. CBF, cerebral blood flow (mL/100g/min); EPI, Echo-planar imaging; sCoV, Spatial Coefficient of Variation.

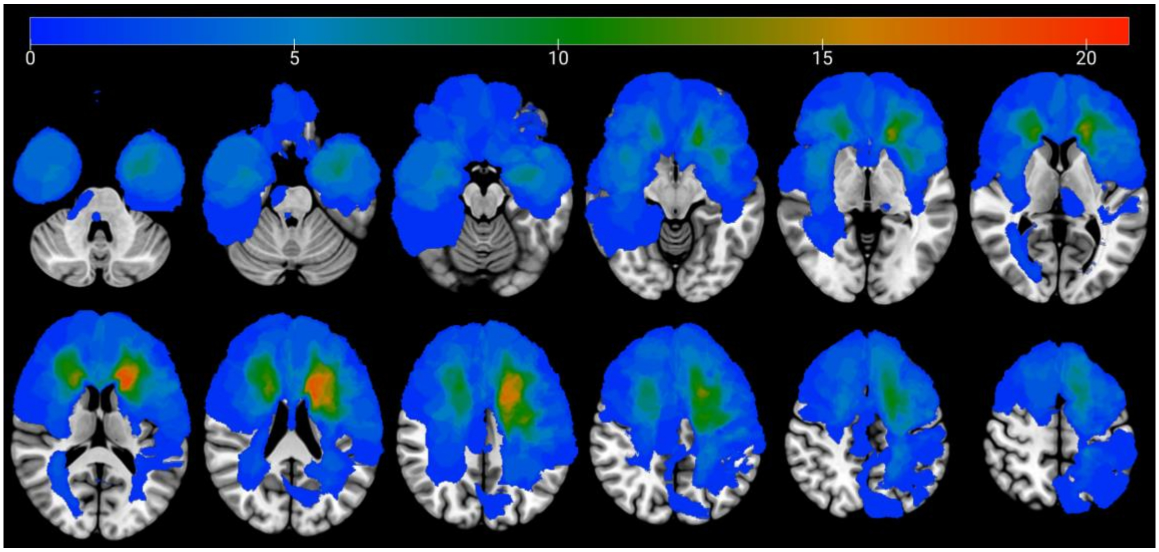

**Figure S2. Lesion distribution heatmap**
Voxelwise lesion frequency across patients included in this study (*n*=44) (peak overlap in frontal regions).

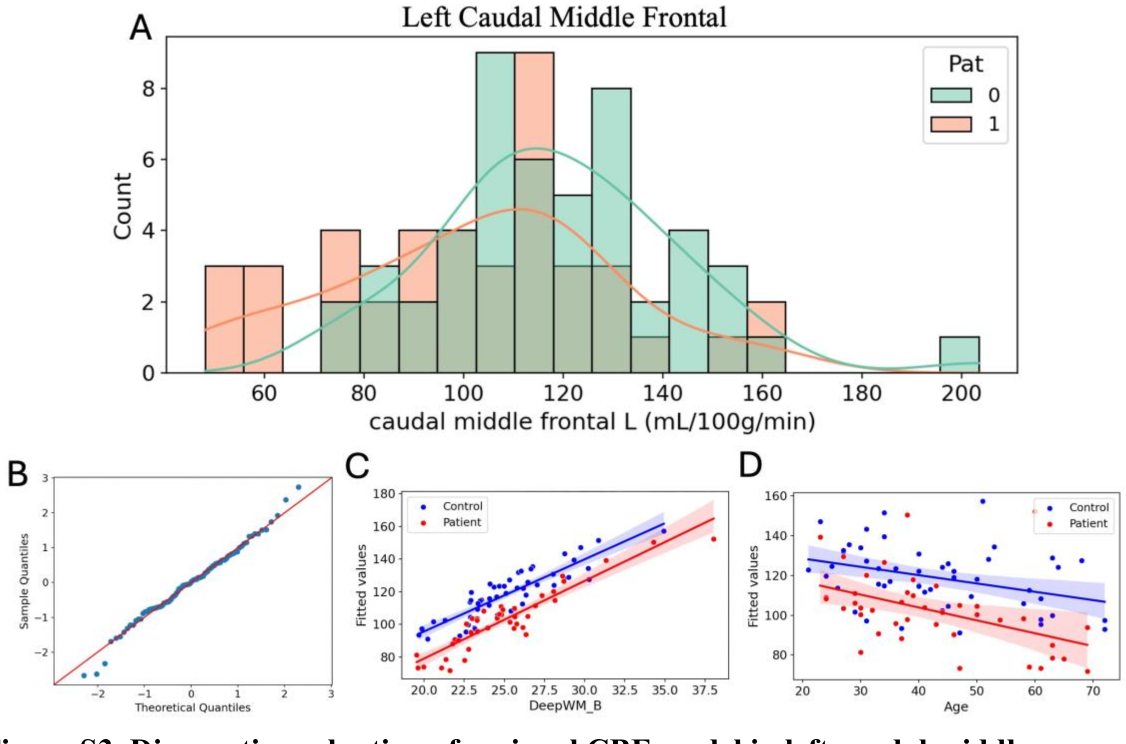

**Figure S3. Diagnostic evaluation of regional CBF model in left caudal middle frontal.**
(A) Distribution plots showing CBF values in patients (orange) and controls (green). (B) Quantile-quantile plot of model residuals demonstrating approximate normality, with points aligning closely to the theoretical reference line. (C) Fitted CBF values and 95% confidence intervals plotted against age of patients (red) and controls (blue), showing a linear association without curvature or violations of model assumptions. (D) Fitted CBF values and 95% confidence intervals plotted against DeepWM CBF of patients (red) and

controls (blue), confirming the expected linear scaling relationship and absence of non-linear patterns. CBF, cerebral blood flow (mL/100g/min); DeepWM, deep white matter.

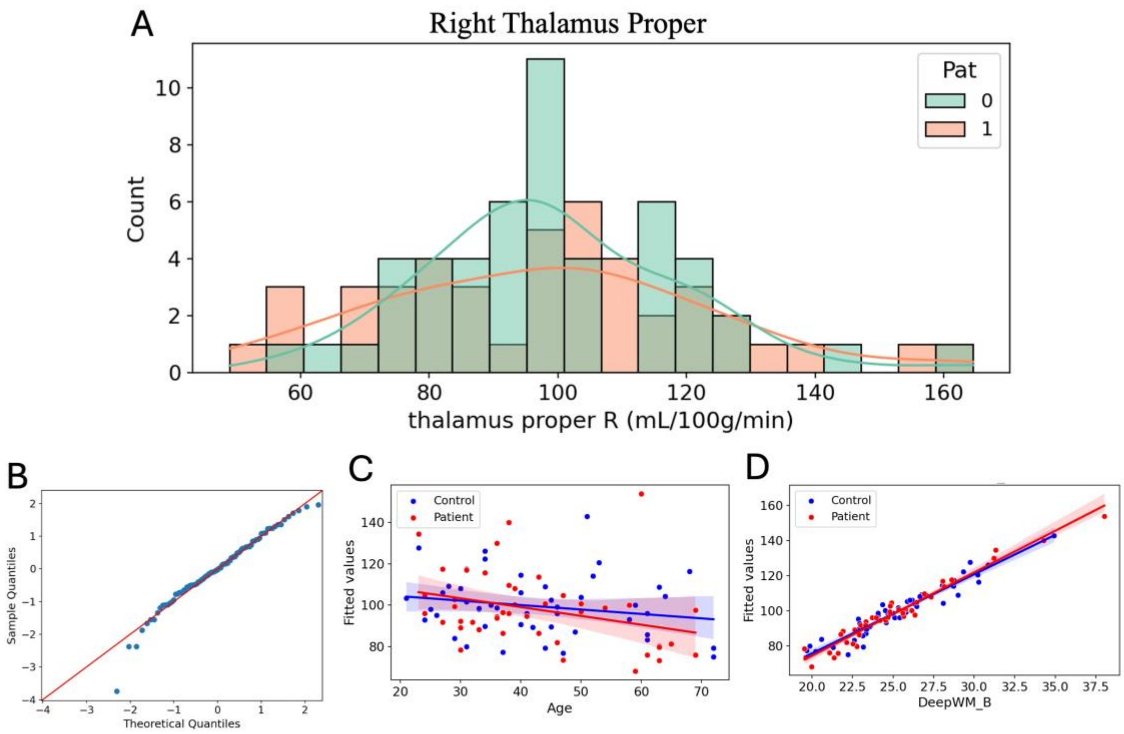

**Figure S4. Diagnostic evaluation of regional CBF model in left thalamus proper.** (A) Distribution plots showing raw CBF values in patients (orange) and controls (green). (B) Quantile-quantile plot of residuals indicating acceptable normality with no systematic departures from the reference line. (C) Fitted CBF values and 95% confidence intervals plotted against age of patients (red) and controls (blue), demonstrating appropriate linearity of this covariate across subjects. (D) Fitted CBF values and 95% confidence intervals plotted against DeepWM CBF of patients (red) and controls (blue), illustrating the expected monotonic relationship and confirming that DeepWM behaves as a stable global-scaling covariate. CBF, cerebral blood flow (mL/100g/min); DeepWM, deep white matter.

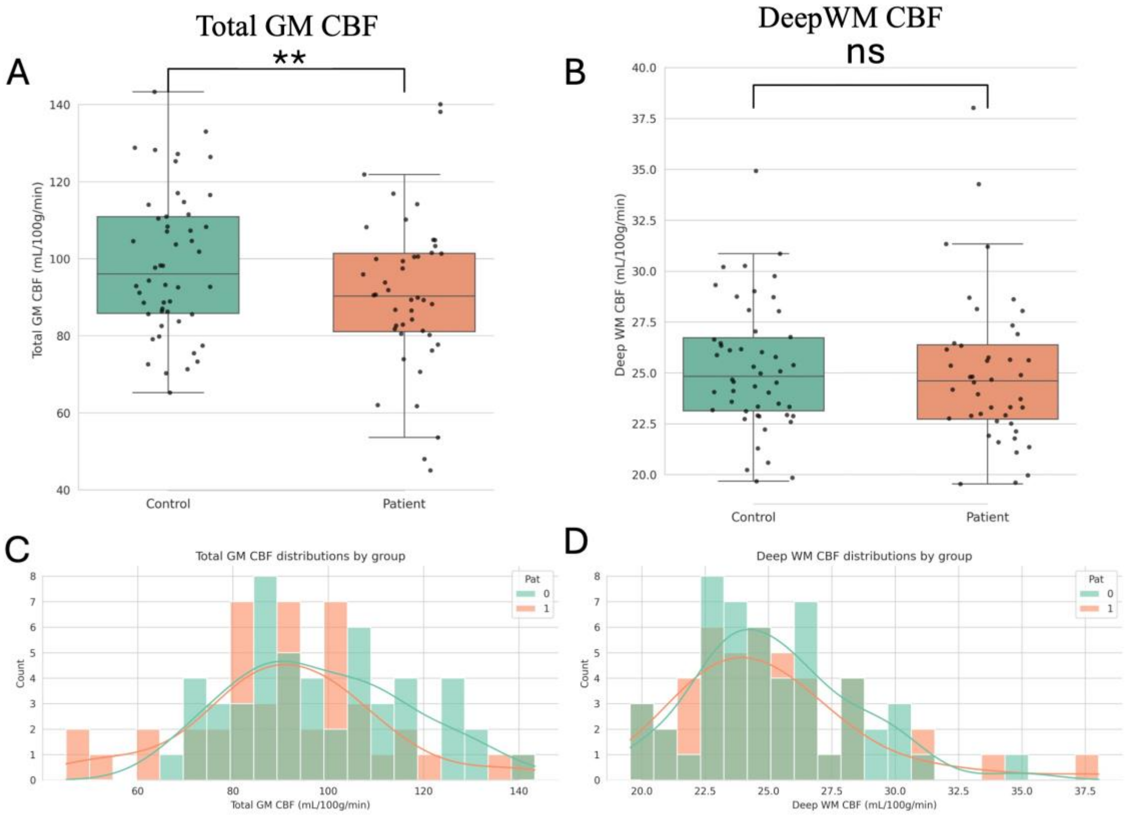

**Figure S5. Group-level distributions of global perfusion indices.**
(A+C) Whole-brain GM CBF was significantly lower in patients (orange) than controls (green) after adjusting for age, sex, and DeepWM CBF in an ANCOVA, the group effect reached significance ($p = 0.006$). (B+D) DeepWM CBF did not differ between patients (orange) and controls (green) after adjusting for age and sex in an ANCOVA ($p = 0.713$), supporting its use as an empirical scaling covariate in this cohort.

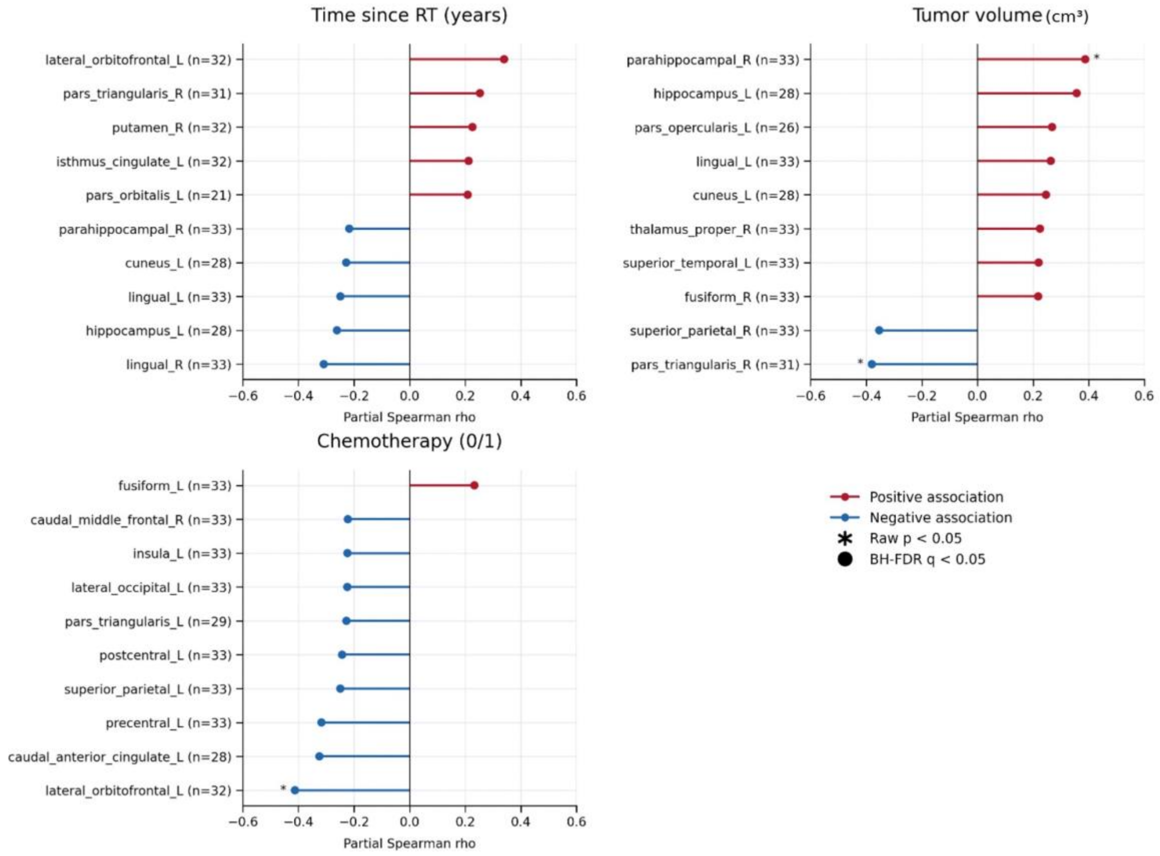

**Figure S6. Partial Spearman associations between regional DKT perfusion and clinical variables after adjustment for age, sex, and DeepWM CBF.**
Panels summarize the top 10 strongest absolute partial Spearman correlations for each clinical variable tested: time since RT (years), tumor volume (cm3), and chemotherapy. Each point represents the adjusted partial Spearman correlation coefficient (ρ) between regional cerebral blood flow (CBF, mL/100g/min) and the corresponding clinical variable for a unilateral DKT region with at least 10 available subjects. Horizontal lines connect each ROI to zero, facilitating visualization of effect direction and magnitude. Red markers indicate positive associations, and blue markers indicate negative associations. ROIs are ordered on partial Spearman correlation strength. Asterisks (*) denote nominal raw $p < 0.05$. No ROI-level association survived Benjamini-Hochberg false discovery rate correction within predictor ($q < 0.05$). Region labels follow unilateral DKT nomenclature (L/R). CBF, cerebral blood flow; DeepWM, deep white matter; DKT, Desikan-Killiany-Tourville; FDR, false discovery rate; ρ, Spearman correlation coefficient; q, Benjamini-Hochberg adjusted p-value.

### 3. Supplementary Tables

**Table S1. Descriptive characteristics of the participants**

| Characteristics | Patients (n=44) | Irradiated patients (n=33, 75%) | Healthy controls (n=50) |
|---|---|---|---|
| **Demographics** | | | |
| **Age at diagnosis (in years)** | | | |
| ***Mean* (SD)** | **34.70 (12.05)** | **35.61 (11.60)** | |
| ***Median* (range)** | **34 (14-63)** | **34 (18-62)** | |
| **Age at inclusion (in years)** | | | |
| ***Mean* (SD)** | **41.27 (12.64)** | **42.39 (12.74)** | **42.42 (13.08)** |
| ***Median* (range)** | **38 (23-69)** | **39 (23-69)** | **40.5 (21-72)** |
| **Sex: females, *n* (%)** | **23 (52)** | **19 (58)** | **25 (50)** |
| **Handedness** | | | |
| **Right, *n (%)*** | **38 (86)** | **28 (85)** | **48 (96)** |
| **Left, *n (%)*** | **4 (9)** | **4 (12)** | **2 (4)** |
| **Both, *n (%)*** | **2 (5)** | **1 (3)** | **0 (0)** |
| **Anti-epileptic drug use** | | | |
| **Yes, *n (%)*** | **29 (66)** | **22 (67)** | **0 (0)** |
| **Monotherapy** | **19** | **13** | **0** |
| **Dual therapy** | **7** | **6** | **0** |
| **Triple therapy** | **3** | **3** | **0** |
| **Tumor treatment** | | | |
| **Time since radiotherapy in years, *mean* (SD)** | | **5.36 (3.26)** | |
| **Time since radiotherapy in years, *median* (range)** | | **4.7 (1.9-16.0)** | |

| | | |
|---|---|---|
| Surgery, *n* (%) | 38 (86) | 27 (82) |
| Total excision | 21 (55) | 12 (44) |
| Subtotal excision | 17 (45) | 15 (56) |
| Biopsy only (no surgery) | 6 (14) | 6 (18) |
| Radiotherapy | | |
| Total dose, *n* (%) | | |
| 54 Gy | | 17 (52) |
| 59.4 Gy | | 6 (18) |
| 60 Gy | | 10 (30) |
| Technique, *n* (%) | | |
| 3DCRT | | 14 (42) |
| VMAT | | 19 (58) |
| Chemotherapy | 30 (68) | 30 (91) |
| PCV, *n* (%) | 20 (67) | 20 (67) |
| TMZ, *n* (%) | 10 (33) | 10 (33) |
| Tumor location* | | |
| Frontal, *n* (%) | 31 (70) | 25 (76) |
| Parietal, *n* (%) | 6 (14) | 5 (15) |
| Temporal, *n* (%) | 12 (27) | 8 (24) |
| Occipital, *n* (%) | 1 (2) | 1 (3) |
| Brainstem, *n* (%) | 2 (5) | 2 (6) |
| Thalamus, *n* (%) | 1 (2) | 1 (3) |
| Involved hemisphere | | |
| Left, *n* (%) | 28 (64) | 22 (67) |

| | | |
|---|---|---|
| **Right, *n* (%)** | **15 (34)** | **10 (30)** |
| **Both, *n* (%)** | **1 (2)** | **1 (3)** |
| **Tumor characteristics** | | |
| **Histology (WHO 2016)** | | |
| **Oligodendroglioma, *n* (%)** | **27 (59)** | **16 (49)** |
| **Diffuse astrocytoma, *n* (%)** | **14 (32)** | **13 (39)** |
| **Anaplastic astrocytoma, *n* (%)** | **2 (5)** | **2 (6)** |
| **Anaplastic oligodendroglioma, *n* (%)** | **2 (5)** | **2 (6)** |
| **WHO grade (2016)** | | |
| **2, *n* (%)** | **35 (80)** | **24 (73)** |
| **3, *n* (%)** | **9 (20)** | **9 (27)** |
| **IDH mutation** | | |
| **IDH1-mutation, *n* (%)** | **30 (68)** | **22 (67)** |
| **IDH1- wild type, *n* (%)** | **9 (20)** | **7 (21)** |
| **IDH1, NOS, *n* (%)** | **5 (11)** | **4 (12)** |

**Caption**: * Overlap of multiple brain tumor locations possible. Demographic, clinical, and treatment characteristics for all included participants. Values are reported as mean (SD), median (range), or n (%) according to variable type. 3DCRT, three-dimensional conformal radiotherapy; Gy, gray; IDH, isocitrate dehydrogenase; NOS, not otherwise specified; PCV, procarbazine lomustine vincristine; TMZ, temozolomide; VMAT, volumetric modulated arc therapy; WHO, World Health Organization.

**Table S2. Neurocognitive tests used for cognitive domain composites and proxy IQ**

| **Cognitive domain** | **Neurocognitive test** | **Outcome measurement** |
|---|---|---|
| Memory | HVLT-R immediate recall | Sum score - learning |
| | HVLT-R delayed recall | Sum score |
| | HVLT-R recognition | Good recognition-mistakes |
| Executive functioning | TMT B | time |

| | | |
|---|---|---|
| | SCWT interference | Interference score |
| | WAIS IV digit span backwards | Total number of series |
| | WAIS IV sequencing | Total number of series |
| Attention / processing speed | WAIS IV symbol substitution | Sum score |
| | TMT A | time |
| | SCWT colors | time |
| | SCWT words | time |
| | WAIS IV digit span forward | Total number of series |
| Motor function | Grooved pegboard | Time (non)dominant hand |
| Proxy IQ | WAIS IV matrix reasoning | Items correct |
| Language | COWAT semantic | Sum of words |
| | COWAT phonemic | Sum of words |

**Caption**: This table summarizes all neurocognitive (sub)tests grouped per domain, with specific outcome metrics used to derive standardized w-scores applied in cognition-CBF analyses.[11] COWAT, Controlled Oral Word Association Test; HVLT-R, Hopkins Verbal Learning Test-Revised; SCWT, Stroop Color-Word Test; TMT, Trail Making Test; WAIS-IV, Wechsler Adult Intelligence Scale-Fourth Edition.

**Table S3. Summary of $M_0$ calibration strategies and outputs**

| **Strategy** | **CSF mask details** | **$M_{0,CSF}$ (a.u.)** | **Exp factor (a.u.)** | **$\lambda_{CSF}$ (mL/g)** | **$M_{0,a}$ (a.u.)** |
|---|---|---|---|---|---|
| Probability map (pCSF ≥ 0.90) | CSF probability map threshold 0.90 | 62,186 | 0.790 | 1.15 | 42,729 |
| Ventricular mask | FastSurfer ventricular CSF | 89,110 | 0.790 | 1.15 | 61,229 |
| Intersection (ventricular ∩ pCSF ≥ 0.90) | Ventricular AND probability ≥0.90 | 89,422 | 0.790 | 1.15 | 61,443 |

**Caption**: Estimated arterial equilibrium magnetization ($M_{0,a}$) using three CSF sampling strategies: probabilistic CSF mask, ventricular mask, and their intersection. All estimates use the reference subject's non-background suppressed M0 control volume with fixed $T_2^*$ correction and water partition coefficient. CSF, cerebrospinal fluid; M0, equilibrium magnetization; $M_{0,a}$, arterial equilibrium magnetization; pCSF, CSF probability map; a.u., arbitrary units; $\lambda_{CSF}$ brain/blood partition coefficient.

**Table S4. Full T2* sensitivity grid for arterial $M_0$**

| $T^*_{2,CSF}$ (ms) | $T^*_{2,a}$ (ms) | exp factor (a.u.) | $M_{0,a}$ (a.u.) |
|---|---|---|---|
| 400 | 40 | 0.754 | 58,640 |
| 400 | 45 | 0.781 | 60,718 |
| 400 | 50 | 0.803 | 62,434 |
| 400 | 55 | 0.822 | 63,874 |
| 400 | 60 | 0.837 | 65,100 |
| 450 | 40 | 0.752 | 58,435 |
| 450 | 45 | 0.778 | 60,507 |
| 450 | 50 | 0.800 | 62,217 |
| 450 | 55 | 0.819 | 63,652 |
| 450 | 60 | 0.834 | 64,873 |
| 500 | 40 | 0.749 | 58,273 |
| 500 | 45 | 0.776 | 60,339 |
| 500 | 50 | 0.798 | 62,044 |
| 500 | 55 | 0.816 | 63,475 |
| 500 | 60 | 0.832 | 64,693 |
| 550 | 40 | 0.748 | 58,140 |
| 550 | 45 | 0.774 | 60,201 |
| 550 | 50 | 0.796 | 61,903 |
| 550 | 55 | 0.814 | 63,331 |
| 550 | 60 | 0.830 | 64,546 |
| 600 | 40 | 0.746 | 58,030 |
| 600 | 45 | 0.773 | 60,087 |
| 600 | 50 | 0.795 | 61,785 |
| 600 | 55 | 0.813 | 63,210 |
| 600 | 60 | 0.829 | 64,423 |
| 650 | 40 | 0.745 | 57,936 |
| 650 | 45 | 0.772 | 59,990 |
| 650 | 50 | 0.793 | 61,686 |
| 650 | 55 | 0.812 | 63,109 |
| 650 | 60 | 0.827 | 64,320 |
| 700 | 40 | 0.744 | 57,857 |
| 700 | 45 | 0.770 | 59,908 |
| 700 | 50 | 0.792 | 61,601 |
| 700 | 55 | 0.810 | 63,022 |
| 700 | 60 | 0.826 | 64,231 |

**Caption**: Complete grid of arterial $M_{0,a}$ estimates across CSF and arterial $T_2^*$ combinations. Values reflect the ventricular ∩ pCSF strategy and show bounded variation across plausible $T_2^*$ ranges. $M_{0,a}$, arterial equilibrium magnetization; exp factor, exponential correction factor; CSF, cerebrospinal fluid.

**Table S5. Global perfusion comparison between patients and controls**

| Measure | Control (n=50) Mean ± SD | Patient (n=44) Mean ± SD | ANCOVA p (group) | partial $\eta^2$ |
|---|---|---|---|---|
| Total GM CBF (mL/100g/min) | 98.6 ± 18.5 | 90.9 ± 20.1 | 0.006 | 0.082 |
| DeepWM CBF (mL/100g/min) | 25.3 ± 3.1 | 25.0 ± 3.7 | 0.713 | 0.002 |

**Caption**: Whole-brain GM CBF and DeepWM CBF were compared between groups using ANCOVA. The Total GM CBF model included age, sex, and DeepWM CBF as covariates, whereas the DeepWM CBF model included age and sex as covariates. DeepWM CBF showed no group difference and was therefore used as an empirical scaling covariate in regional perfusion analyses. ANCOVA, analysis of covariance; CBF, cerebral blood flow; DeepWM, deep white matter; GM, gray matter.

**Table S6. DKT regions excluded due to insufficient effective volume**

| Region | Controls Available (n) | Patients Available (n) | Reason for Exclusion |
|---|---|---|---|
| pallidum_L | 0 | 0 | Did not meet ≥20 participants per group with ≥1 $mm^3$ usable tissue |
| pallidum_R | 0 | 0 | Did not meet ≥20 participants per group with ≥1 $mm^3$ usable tissue |
| amygdala_L | 0 | 0 | Did not meet ≥20 participants per group with ≥1 $mm^3$ usable tissue |
| amygdala_R | 0 | 0 | Did not meet ≥20 participants per group with ≥1 $mm^3$ usable tissue |
| accumbens_area_L | 0 | 0 | Did not meet ≥20 participants per group with ≥1 $mm^3$ usable tissue |

| | | | |
|---|---|---|---|
| accumbens_area_R | 0 | 0 | Did not meet ≥20 participants per group with ≥1 mm³ usable tissue |
| ventral_DC_L | 0 | 0 | Did not meet ≥20 participants per group with ≥1 mm³ usable tissue |
| ventral_DC_R | 1 | 0 | Did not meet ≥20 participants per group with ≥1 mm³ usable tissue |
| basal_forebrain_L | 0 | 0 | Did not meet ≥20 participants per group with ≥1 mm³ usable tissue |
| basal_forebrain_R | 0 | 0 | Did not meet ≥20 participants per group with ≥1 mm³ usable tissue |
| pericalcarine_L | 22 | 10 | Did not meet ≥20 participants per group with ≥1 mm³ usable tissue |
| pericalcarine_R | 17 | 8 | Did not meet ≥20 participants per group with ≥1 mm³ usable tissue |
| transverse_temporal_L | 9 | 3 | Did not meet ≥20 participants per group with ≥1 mm³ usable tissue |
| transverse_temporal_R | 4 | 1 | Did not meet ≥20 participants per group with ≥1 mm³ usable tissue |

**Caption**: ROIs were excluded from regional CBF analyses when lesion masking and atlas intersection did not meet inclusion criterion of ≥20 participants per group with ≥1 mm³ usable tissue. Availability counts are reported as n and denote the number of subjects with sufficient effective ROI volume after lesion masking and atlas intersection. Counts vary by ROI because lesion extent and ROI overlap differed across subjects. DKT, Desikan-Killiany-Tourville atlas; ROI, region of interest.

**Table S7. DKT regions included in the primary perfusion analysis**

| Region | Controls Available (n) | Patients Available (n) | Adjusted mean - Control (95% CI) | Adjusted mean - Patient (95% CI) | Partial η² (group) | ANCOVA p (group) | FDR q (BH) | FDR significant (q<0.01) |
|---|---|---|---|---|---|---|---|---|
| thalamus_proper_L | 50 | 44 | 98.4 (92.4-104.3) | 100.9 (94.8-107.0) | 0.006 | 0.466 | 0.496 | No |
| thalamus_proper_R | 50 | 44 | 100.7 (95.0-106.3) | 100.2 (94.4-106.0) | 0.000 | 0.892 | 0.892 | No |
| caudate_L | 50 | 40 | 87.2 (80.9-93.4) | 84.2 (77.4-90.9) | 0.007 | 0.432 | 0.467 | No |
| caudate_R | 50 | 42 | 83.5 (78.3-88.8) | 80.4 (74.8-86.0) | 0.011 | 0.324 | 0.382 | No |
| putamen_L | 50 | 42 | 69.4 (64.6-74.1) | 70.2 (65.1-75.2) | 0.001 | 0.775 | 0.787 | No |
| putamen_R | 50 | 43 | 68.9 (64.3-73.6) | 67.7 (62.9-72.6) | 0.002 | 0.667 | 0.687 | No |
| hippocampus_L | 50 | 39 | 80.3 (75.0-85.6) | 77.4 (71.6-83.1) | 0.009 | 0.375 | 0.422 | No |
| hippocampus_R | 50 | 42 | 79.6 (74.3-84.9) | 70.4 (64.9-76.0) | 0.088 | 0.005 | 0.023 | No |
| caudal_anterior_cingulate_L | 50 | 38 | 105.5 (99.7-111.3) | 99.1 (92.9-105.3) | 0.039 | 0.070 | 0.107 | No |
| caudal_anterior_cingulate_R | 50 | 38 | 107.3 (101.3-113.3) | 103.2 (96.7-109.7) | 0.015 | 0.269 | 0.328 | No |
| caudal_middle_frontal_L | 50 | 42 | 115.8 (108.8-122.9) | 100.8 (93.5-108.1) | 0.128 | <0.001 | 0.007 | Yes |
| caudal_middle_frontal_R | 50 | 43 | 116.6 (109.8-123.3) | 106.7 (99.8-113.7) | 0.064 | 0.016 | 0.039 | No |
| cuneus_L | 50 | 39 | 89.4 (82.0-96.8) | 82.4 (74.5-90.2) | 0.028 | 0.121 | 0.171 | No |
| cuneus_R | 50 | 44 | 90.1 (83.3-96.8) | 85.5 (78.5-92.4) | 0.015 | 0.253 | 0.321 | No |
| entorhinal_L | 50 | 41 | 60.8 (56.2-65.4) | 57.3 (52.4-62.1) | 0.019 | 0.196 | 0.264 | No |

| | | | | | | | | |
|---|---|---|---|---|---|---|---|---|
| entorhinal_R | 50 | 41 | 59.2 (55.0-63.4) | 52.9 (48.4-57.3) | 0.068 | 0.015 | 0.039 | No |
| fusiform_L | 50 | 44 | 95.0 (90.2-99.9) | 89.1 (84.1-94.1) | 0.046 | 0.041 | 0.074 | No |
| fusiform_R | 50 | 44 | 98.6 (93.2-104.1) | 92.4 (86.8-98.0) | 0.040 | 0.056 | 0.091 | No |
| inferior_parietal_L | 50 | 44 | 111.7 (104.7-118.6) | 96.7 (89.6-103.8) | 0.130 | <0.001 | 0.007 | Yes |
| inferior_parietal_R | 50 | 44 | 118.8 (112.1-125.5) | 105.6 (98.7-112.5) | 0.110 | 0.001 | 0.013 | No |
| inferior_temporal_L | 50 | 44 | 71.1 (66.7-75.5) | 64.1 (59.6-68.6) | 0.075 | 0.008 | 0.029 | No |
| inferior_temporal_R | 50 | 44 | 75.1 (69.8-80.4) | 67.1 (61.7-72.5) | 0.069 | 0.012 | 0.037 | No |
| isthmus_cingulate_L | 50 | 43 | 124.8 (118.5-131.1) | 115.7 (109.3-122.1) | 0.063 | 0.017 | 0.039 | No |
| isthmus_cingulate_R | 49 | 43 | 130.8 (123.9-137.6) | 122.6 (115.3-129.9) | 0.044 | 0.049 | 0.083 | No |
| lateral_occipital_L | 50 | 44 | 73.5 (69.0-78.1) | 67.2 (62.5-71.8) | 0.059 | 0.020 | 0.044 | No |
| lateral_occipital_R | 50 | 44 | 74.3 (69.5-79.1) | 68.5 (63.6-73.4) | 0.045 | 0.044 | 0.076 | No |
| lateral_orbitofrontal_L | 50 | 43 | 95.3 (89.9-100.6) | 88.7 (83.1-94.2) | 0.046 | 0.041 | 0.074 | No |
| lateral_orbitofrontal_R | 50 | 44 | 96.0 (91.0-101.0) | 87.6 (82.5-92.8) | 0.082 | 0.006 | 0.024 | No |
| lingual_L | 50 | 44 | 106.3 (100.8-111.8) | 103.4 (97.7-109.0) | 0.009 | 0.370 | 0.422 | No |
| lingual_R | 50 | 44 | 105.5 (99.7-111.4) | 101.8 (95.7-107.8) | 0.013 | 0.281 | 0.337 | No |
| medial_orbitofrontal_L | 50 | 43 | 82.9 (78.4-87.4) | 75.5 (70.9-80.2) | 0.080 | 0.007 | 0.026 | No |
| medial_orbitofrontal_R | 50 | 42 | 78.3 (73.5-83.1) | 75.8 (70.7-80.9) | 0.008 | 0.396 | 0.435 | No |

| | | | | | | | | |
|---|---|---|---|---|---|---|---|---|
| middle_temporal_L | 50 | 44 | 99.3 (93.8-104.7) | 89.4 (83.8-94.9) | 0.097 | 0.003 | 0.020 | No |
| middle_temporal_R | 50 | 44 | 97.9 (92.7-103.1) | 89.3 (84.0-94.6) | 0.082 | 0.006 | 0.024 | No |
| parahippocampal_L | 50 | 43 | 99.1 (92.8-105.5) | 93.1 (86.4-99.7) | 0.028 | 0.113 | 0.162 | No |
| parahippocampal_R | 50 | 43 | 97.2 (90.8-103.6) | 90.7 (84.2-97.3) | 0.032 | 0.091 | 0.133 | No |
| paracentral_L | 50 | 44 | 100.3 (94.6-106.0) | 96.5 (90.7-102.3) | 0.014 | 0.257 | 0.321 | No |
| paracentral_R | 50 | 44 | 107.1 (101.4-112.7) | 104.8 (99.0-110.5) | 0.005 | 0.489 | 0.512 | No |
| pars_opercularis_L | 50 | 37 | 118.7 (111.3-126.1) | 114.7 (106.7-122.7) | 0.010 | 0.377 | 0.422 | No |
| pars_opercularis_R | 50 | 42 | 124.1 (117.6-130.5) | 112.2 (105.3-119.1) | 0.097 | 0.003 | 0.020 | No |
| pars_orbitalis_L | 48 | 32 | 113.0 (104.9-121.1) | 106.7 (97.5-115.9) | 0.020 | 0.219 | 0.289 | No |
| pars_orbitalis_R | 40 | 34 | 119.2 (111.7-126.7) | 108.1 (100.6-115.6) | 0.083 | 0.015 | 0.039 | No |
| pars_triangularis_L | 50 | 40 | 115.1 (108.0-122.2) | 102.3 (94.7-109.8) | 0.095 | 0.004 | 0.020 | No |
| pars_triangularis_R | 50 | 41 | 118.0 (111.5-124.5) | 106.0 (99.0-113.0) | 0.097 | 0.003 | 0.020 | No |
| postcentral_L | 50 | 44 | 102.6 (97.6-107.5) | 96.0 (90.9-101.1) | 0.054 | 0.027 | 0.056 | No |
| postcentral_R | 50 | 44 | 102.8 (97.7-107.9) | 96.3 (91.1-101.5) | 0.050 | 0.033 | 0.064 | No |
| posterior_cingulate_L | 50 | 44 | 110.3 (104.8-115.9) | 105.5 (99.8-111.1) | 0.025 | 0.137 | 0.188 | No |
| posterior_cingulate_R | 50 | 43 | 116.4 (110.3-122.5) | 112.1 (105.9-118.4) | 0.016 | 0.242 | 0.313 | No |
| precentral_L | 50 | 44 | 113.5 (107.9-119.0) | 103.8 (98.1-109.4) | 0.090 | 0.004 | 0.020 | No |

| | | | | | | | | |
|---|---|---|---|---|---|---|---|---|
| precentral_R | 50 | 44 | 111.3 (105.2-117.3) | 103.5 (97.4-109.7) | 0.050 | 0.032 | 0.064 | No |
| precuneus_L | 50 | 44 | 125.7 (119.3-132.0) | 115.6 (109.1-122.1) | 0.075 | 0.009 | 0.029 | No |
| precuneus_R | 50 | 44 | 132.8 (126.2-139.4) | 121.8 (115.1-128.5) | 0.082 | 0.006 | 0.024 | No |
| rostral_anterior_cingulate_L | 50 | 42 | 101.0 (95.2-106.7) | 94.5 (88.4-100.7) | 0.039 | 0.065 | 0.103 | No |
| rostral_anterior_cingulate_R | 50 | 39 | 96.2 (90.8-101.6) | 89.3 (83.5-95.0) | 0.050 | 0.039 | 0.073 | No |
| rostral_middle_frontal_L | 50 | 42 | 117.5 (110.8-124.2) | 102.0 (94.8-109.1) | 0.147 | <0.001 | 0.007 | Yes |
| rostral_middle_frontal_R | 50 | 44 | 108.4 (102.5-114.3) | 95.7 (89.6-101.8) | 0.128 | <0.001 | 0.007 | Yes |
| superior_frontal_L | 50 | 44 | 104.4 (99.0-109.8) | 94.9 (89.4-100.4) | 0.091 | 0.004 | 0.020 | No |
| superior_frontal_R | 50 | 44 | 103.2 (97.9-108.6) | 95.4 (90.0-100.9) | 0.065 | 0.015 | 0.039 | No |
| superior_parietal_L | 50 | 44 | 119.5 (112.2-126.7) | 103.4 (95.9-110.8) | 0.135 | <0.001 | 0.007 | Yes |
| superior_parietal_R | 50 | 44 | 116.6 (110.8-122.5) | 104.0 (98.0-110.0) | 0.131 | <0.001 | 0.007 | Yes |
| superior_temporal_L | 50 | 44 | 94.5 (89.3-99.6) | 87.6 (82.4-92.9) | 0.054 | 0.027 | 0.056 | No |
| superior_temporal_R | 50 | 44 | 94.6 (89.7-99.6) | 87.5 (82.5-92.6) | 0.063 | 0.017 | 0.039 | No |
| supramarginal_L | 50 | 43 | 110.8 (105.2-116.4) | 102.7 (96.9-108.5) | 0.063 | 0.017 | 0.039 | No |
| supramarginal_R | 50 | 44 | 112.4 (106.4-118.4) | 103.6 (97.5-109.7) | 0.064 | 0.015 | 0.039 | No |
| insula_L | 50 | 44 | 91.4 (86.4-96.5) | 86.2 (81.0-91.4) | 0.033 | 0.085 | 0.127 | No |
| insula_R | 50 | 42 | 94.2 (89.7-98.6) | 89.0 (84.3-93.7) | 0.041 | 0.056 | 0.091 | No |

**Caption**: For each included ROI, model-based adjusted means (95% CI), effect sizes using partial $\eta^2$, and BH-FDR results are reported from ANCOVA models (CBF ~ group + age + sex + DeepWM CBF). Control and patient availability counts are reported as n and denote the number of subjects contributing to each ROI after lesion masking and atlas intersection. Counts vary by ROI because usable effective tissue volume differed across subjects. FDR significance threshold: $q < 0.01$. CBF, cerebral blood flow; CI, confidence interval; DKT, Desikan-Killiany-Tourville atlas; DeepWM, deep white matter; FDR, false discovery rate; ROI, region of interest.

**Table S8. ROI selection based on regional RT dose variability**

| Region | Mean RT dose (Gy) | SD RT dose (Gy) |
|---|---|---|
| thalamus_proper_L | 41.2 | 15.4 |
| thalamus_proper_R | 37.8 | 13.8 |
| caudate_L | 43.8 | 15.2 |
| caudate_R | 40.7 | 15.2 |
| putamen_L | 41.8 | 17.5 |
| putamen_R | 36.4 | 16.2 |
| pallidum_L | 42.3 | 16.8 |
| pallidum_R | 37.6 | 15.9 |
| hippocampus_L | 35.2 | 19.9 |
| hippocampus_R | 27.8 | 18.5 |
| amygdala_L | 37.2 | 19.4 |
| amygdala_R | 30.8 | 19.0 |
| accumbens_area_L | 43.3 | 15.4 |
| accumbens_area_R | 40.3 | 15.5 |
| basal_forebrain_L | 40.2 | 16.4 |
| basal_forebrain_R | 40.7 | 16.0 |
| caudal_anterior_cingulate_L | 42.9 | 16.1 |
| caudal_anterior_cingulate_R | 39.7 | 16.8 |
| caudal_middle_frontal_L | 31.4 | 21.1 |
| caudal_middle_frontal_R | 25.4 | 17.9 |
| cuneus_L | 15.8 | 14.8 |
| cuneus_R | 14.2 | 14.0 |
| entorhinal_L | 30.8 | 21.7 |
| entorhinal_R | 24.6 | 20.2 |
| fusiform_L | 23.8 | 18.2 |
| fusiform_R | 17.6 | 16.7 |
| inferior_parietal_L | 18.9 | 18.1 |
| inferior_parietal_R | 12.3 | 12.9 |
| inferior_temporal_L | 23.3 | 17.8 |
| inferior_temporal_R | 17.0 | 16.8 |
| isthmus_cingulate_L | 28.0 | 18.3 |
| isthmus_cingulate_R | 25.8 | 16.6 |
| lateral_occipital_L | 14.1 | 13.6 |

| | | |
|---|---|---|
| lateral_occipital_R | 10.7 | 13.3 |
| lateral_orbitofrontal_L | 34.2 | 18.7 |
| lateral_orbitofrontal_R | 29.2 | 16.7 |
| lingual_L | 19.0 | 15.3 |
| lingual_R | 16.9 | 15.2 |
| medial_orbitofrontal_L | 34.9 | 16.9 |
| medial_orbitofrontal_R | 34.4 | 17.0 |
| middle_temporal_L | 26.9 | 18.4 |
| middle_temporal_R | 18.8 | 15.9 |
| parahippocampal_L | 30.9 | 21.1 |
| parahippocampal_R | 24.4 | 19.2 |
| paracentral_L | 22.8 | 19.6 |
| paracentral_R | 20.9 | 16.8 |
| pars_opercularis_L | 38.9 | 19.2 |
| pars_opercularis_R | 31.5 | 17.4 |
| pars_orbitalis_L | 35.7 | 19.2 |
| pars_orbitalis_R | 29.1 | 17.1 |
| pars_triangularis_L | 37.5 | 18.2 |
| pars_triangularis_R | 30.4 | 17.2 |
| pericalcarine_L | 16.6 | 14.9 |
| pericalcarine_R | 13.9 | 14.7 |
| postcentral_L | 26.3 | 17.9 |
| postcentral_R | 20.2 | 13.2 |
| posterior_cingulate_L | 33.1 | 18.1 |
| posterior_cingulate_R | 31.5 | 17.2 |
| precentral_L | 29.4 | 17.8 |
| precentral_R | 22.9 | 14.2 |
| precuneus_L | 20.0 | 18.1 |
| precuneus_R | 17.3 | 15.2 |
| rostral_anterior_cingulate_L | 42.4 | 15.1 |
| rostral_anterior_cingulate_R | 40.2 | 16.9 |
| rostral_middle_frontal_L | 34.4 | 18.3 |
| rostral_middle_frontal_R | 28.8 | 17.5 |
| superior_frontal_L | 31.3 | 16.2 |
| superior_frontal_R | 28.6 | 16.0 |
| superior_parietal_L | 16.1 | 18.1 |
| superior_parietal_R | 12.0 | 12.3 |
| superior_temporal_L | 32.6 | 18.5 |
| superior_temporal_R | 23.9 | 15.6 |
| supramarginal_L | 25.2 | 19.0 |
| supramarginal_R | 19.4 | 15.5 |
| transverse_temporal_L | 37.1 | 20.6 |
| transverse_temporal_R | 27.8 | 16.0 |
| insula_L | 39.7 | 18.6 |
| insula_R | 32.4 | 16.1 |

**Caption**: ROIs meeting the RT-dose variability eligibility criteria (SD ≥10 Gy) included in RT dose-CBF analyses. Values represent cohort-level absorbed RT dose metrics. Gy, gray; ROI, region of interest; SD, standard deviation.

**Table S9. ROI-wise OLS regression of RT dose-CBF associations**

| Region | n | Slope (CBF/Gy) | SE | t | p | q (FDR) | CI95 low | CI95 high |
|---|---|---|---|---|---|---|---|---|
| caudal_anterior_cingulate_L | 28 | -0.02 | 0.22 | -0.09 | 0.929 | 0.980 | -0.47 | 0.43 |
| caudal_anterior_cingulate_R | 29 | 0.21 | 0.24 | 0.87 | 0.391 | 0.859 | -0.29 | 0.71 |
| caudal_middle_frontal_L | 31 | -0.41 | 0.21 | -1.91 | 0.067 | 0.518 | -0.85 | 0.03 |
| caudal_middle_frontal_R | 33 | -0.19 | 0.20 | -0.94 | 0.356 | 0.859 | -0.61 | 0.23 |
| caudate_L | 29 | -0.15 | 0.30 | -0.51 | 0.615 | 0.859 | -0.77 | 0.46 |
| caudate_R | 31 | 0.38 | 0.21 | 1.80 | 0.083 | 0.518 | -0.05 | 0.81 |
| cuneus_L | 28 | 0.43 | 0.38 | 1.14 | 0.265 | 0.781 | -0.35 | 1.22 |
| cuneus_R | 33 | 0.23 | 0.33 | 0.69 | 0.493 | 0.859 | -0.45 | 0.90 |
| entorhinal_L | 30 | 0.10 | 0.11 | 0.92 | 0.368 | 0.859 | -0.12 | 0.32 |
| entorhinal_R | 30 | 0.11 | 0.14 | 0.79 | 0.439 | 0.859 | -0.17 | 0.38 |
| fusiform_L | 33 | 0.13 | 0.17 | 0.74 | 0.467 | 0.859 | -0.22 | 0.48 |
| fusiform_R | 33 | 0.20 | 0.21 | 0.94 | 0.358 | 0.859 | -0.23 | 0.63 |
| hippocampus_L | 28 | -0.02 | 0.17 | -0.12 | 0.906 | 0.980 | -0.36 | 0.33 |
| hippocampus_R | 32 | 0.24 | 0.17 | 1.41 | 0.169 | 0.621 | -0.11 | 0.59 |
| inferior_parietal_L | 33 | 0.00 | 0.24 | 0.00 | 0.997 | 0.997 | -0.48 | 0.48 |
| inferior_parietal_R | 33 | -0.14 | 0.40 | -0.35 | 0.726 | 0.871 | -0.97 | 0.68 |
| inferior_temporal_L | 33 | 0.20 | 0.14 | 1.38 | 0.179 | 0.621 | -0.10 | 0.48 |
| inferior_temporal_R | 33 | 0.42 | 0.20 | 2.08 | 0.046 | 0.518 | 0.01 | 0.83 |
| insula_L | 33 | -0.24 | 0.16 | -1.51 | 0.142 | 0.551 | -0.57 | 0.09 |
| insula_R | 31 | 0.07 | 0.18 | 0.38 | 0.704 | 0.871 | -0.29 | 0.43 |
| isthmus_cingulate_L | 32 | 0.16 | 0.22 | 0.72 | 0.479 | 0.859 | -0.30 | 0.62 |
| isthmus_cingulate_R | 32 | 0.35 | 0.23 | 1.53 | 0.138 | 0.551 | -0.12 | 0.81 |
| lateral_occipital_L | 33 | 0.13 | 0.21 | 0.62 | 0.538 | 0.859 | -0.30 | 0.56 |
| lateral_occipital_R | 33 | -0.34 | 0.21 | -1.61 | 0.120 | 0.546 | -0.78 | 0.09 |
| lateral_orbitofrontal_L | 32 | -0.23 | 0.17 | -1.33 | 0.193 | 0.621 | -0.58 | 0.12 |
| lateral_orbitofrontal_R | 33 | -0.04 | 0.19 | -0.24 | 0.813 | 0.926 | -0.43 | 0.34 |
| lingual_L | 33 | 0.12 | 0.22 | 0.54 | 0.591 | 0.859 | -0.33 | 0.56 |
| lingual_R | 33 | 0.49 | 0.25 | 1.99 | 0.057 | 0.518 | -0.01 | 1.00 |
| medial_orbitofrontal_L | 32 | -0.26 | 0.14 | -1.78 | 0.086 | 0.518 | -0.56 | 0.04 |
| medial_orbitofrontal_R | 32 | -0.02 | 0.14 | -0.16 | 0.876 | 0.963 | -0.32 | 0.27 |
| middle_temporal_L | 33 | 0.07 | 0.17 | 0.38 | 0.709 | 0.871 | -0.29 | 0.42 |
| middle_temporal_R | 33 | 0.22 | 0.23 | 1.00 | 0.328 | 0.859 | -0.24 | 0.68 |
| paracentral_L | 33 | 0.09 | 0.19 | 0.49 | 0.625 | 0.859 | -0.29 | 0.48 |
| paracentral_R | 33 | -0.01 | 0.22 | -0.04 | 0.970 | 0.985 | -0.46 | 0.44 |
| parahippocampal_L | 32 | 0.07 | 0.18 | 0.40 | 0.694 | 0.871 | -0.29 | 0.43 |
| parahippocampal_R | 33 | 0.32 | 0.20 | 1.58 | 0.124 | 0.546 | -0.09 | 0.74 |
| pars_opercularis_L | 26 | -0.14 | 0.29 | -0.46 | 0.649 | 0.871 | -0.75 | 0.48 |
| pars_opercularis_R | 31 | 0.12 | 0.22 | 0.56 | 0.583 | 0.859 | -0.33 | 0.57 |
| pars_orbitalis_L | 21 | -0.30 | 0.44 | -0.68 | 0.509 | 0.859 | -1.24 | 0.64 |
| pars_orbitalis_R | 24 | -0.12 | 0.31 | -0.41 | 0.690 | 0.871 | -0.77 | 0.52 |
| pars_triangularis_L | 29 | -0.34 | 0.26 | -1.32 | 0.198 | 0.621 | -0.87 | 0.19 |
| pars_triangularis_R | 31 | 0.06 | 0.25 | 0.25 | 0.804 | 0.926 | -0.46 | 0.59 |

| | | | | | | | | |
|---|---|---|---|---|---|---|---|---|
| postcentral_L | 33 | -0.30 | 0.16 | -1.87 | 0.072 | 0.518 | -0.64 | 0.03 |
| postcentral_R | 33 | -0.14 | 0.23 | -0.61 | 0.547 | 0.859 | -0.60 | 0.33 |
| posterior_cingulate_L | 33 | 0.07 | 0.18 | 0.37 | 0.713 | 0.871 | -0.31 | 0.44 |
| posterior_cingulate_R | 32 | 0.21 | 0.19 | 1.12 | 0.272 | 0.781 | -0.18 | 0.60 |
| precentral_L | 33 | -0.50 | 0.18 | -2.76 | 0.010 | 0.221 | -0.87 | -0.13 |
| precentral_R | 33 | -0.02 | 0.25 | -0.08 | 0.935 | 0.980 | -0.53 | 0.48 |
| precuneus_L | 33 | 0.06 | 0.20 | 0.31 | 0.761 | 0.897 | -0.35 | 0.47 |
| precuneus_R | 33 | 0.12 | 0.24 | 0.50 | 0.620 | 0.859 | -0.36 | 0.60 |
| putamen_L | 31 | -0.10 | 0.18 | -0.59 | 0.562 | 0.859 | -0.47 | 0.26 |
| putamen_R | 32 | 0.32 | 0.17 | 1.85 | 0.076 | 0.518 | -0.04 | 0.68 |
| rostral_anterior_cingulate_L | 31 | -0.20 | 0.24 | -0.83 | 0.412 | 0.859 | -0.69 | 0.29 |
| rostral_anterior_cingulate_R | 29 | 0.13 | 0.19 | 0.69 | 0.497 | 0.859 | -0.27 | 0.53 |
| rostral_middle_frontal_L | 31 | -0.46 | 0.25 | -1.84 | 0.078 | 0.518 | -0.98 | 0.06 |
| rostral_middle_frontal_R | 33 | -0.17 | 0.19 | -0.89 | 0.384 | 0.859 | -0.56 | 0.22 |
| superior_frontal_L | 33 | -0.30 | 0.18 | -1.62 | 0.117 | 0.546 | -0.68 | 0.08 |
| superior_frontal_R | 33 | -0.10 | 0.17 | -0.57 | 0.575 | 0.859 | -0.46 | 0.26 |
| superior_parietal_L | 33 | -0.18 | 0.23 | -0.78 | 0.442 | 0.859 | -0.64 | 0.29 |
| superior_parietal_R | 33 | 0.01 | 0.23 | 0.04 | 0.969 | 0.985 | -0.45 | 0.47 |
| superior_temporal_L | 33 | 0.03 | 0.16 | 0.16 | 0.875 | 0.963 | -0.30 | 0.35 |
| superior_temporal_R | 33 | 0.51 | 0.18 | 2.84 | 0.008 | 0.221 | 0.14 | 0.87 |
| supramarginal_L | 32 | -0.09 | 0.17 | -0.54 | 0.594 | 0.859 | -0.44 | 0.26 |
| supramarginal_R | 33 | 0.16 | 0.26 | 0.59 | 0.560 | 0.859 | -0.39 | 0.70 |
| thalamus_proper_L | 33 | 0.40 | 0.23 | 1.71 | 0.098 | 0.540 | -0.08 | 0.87 |
| thalamus_proper_R | 33 | 0.68 | 0.20 | 3.45 | 0.002 | 0.119 | 0.28 | 1.08 |

**Caption**: Ordinary least-squares regression estimates for the association between local RT dose and regional CBF across selected ROIs, adjusted for age, sex, and DeepWM CBF. N denotes the number of irradiated patients contributing to each ROI-wise regression. Counts may vary across ROIs because inclusion additionally required RT dose eligibility and sufficient usable ROI volume after lesion masking and atlas intersection. Includes slope, SE, t statistic, exact p-value, FDR-adjusted q-value, and 95% CI. CBF, cerebral blood flow; CI, confidence interval; DeepWM, deep white matter; FDR, false discovery rate; ROI, region of interest.

**Table S10. ROI-wise sample sizes and eligibility for the three-group RT dose comparison**

| ROI | n control | n low-RT dose | n high-RT dose | Eligible for ANCOVA |
|---|---|---|---|---|
| thalamus_proper_L | 50 | 2 | 19 | No |
| thalamus_proper_R | 50 | 3 | 16 | No |
| caudate_L | 50 | 1 | 19 | No |
| caudate_R | 50 | 2 | 15 | No |
| putamen_L | 50 | 3 | 18 | No |
| putamen_R | 50 | 4 | 11 | No |
| hippocampus_L | 50 | 7 | 10 | No |
| hippocampus_R | 50 | 11 | 8 | Yes |
| caudal_anterior_cingulate_L | 50 | 1 | 18 | No |

| | | | | |
|---|---|---|---|---|
| caudal_anterior_cingulate_R | 50 | 3 | 15 | No |
| caudal_middle_frontal_L | 50 | 10 | 11 | Yes |
| caudal_middle_frontal_R | 50 | 10 | 8 | Yes |
| cuneus_L | 50 | 17 | 1 | No |
| cuneus_R | 50 | 23 | 1 | No |
| entorhinal_L | 50 | 12 | 11 | Yes |
| entorhinal_R | 50 | 16 | 5 | No |
| fusiform_L | 50 | 16 | 8 | Yes |
| fusiform_R | 50 | 20 | 5 | No |
| inferior_parietal_L | 50 | 19 | 6 | No |
| inferior_parietal_R | 50 | 24 | 0 | No |
| inferior_temporal_L | 50 | 15 | 9 | Yes |
| inferior_temporal_R | 50 | 20 | 3 | No |
| isthmus_cingulate_L | 50 | 8 | 8 | Yes |
| isthmus_cingulate_R | 49 | 10 | 7 | No |
| lateral_occipital_L | 50 | 23 | 2 | No |
| lateral_occipital_R | 50 | 27 | 1 | No |
| lateral_orbitofrontal_L | 50 | 6 | 15 | No |
| lateral_orbitofrontal_R | 50 | 7 | 11 | No |
| lingual_L | 50 | 17 | 4 | No |
| lingual_R | 50 | 19 | 2 | No |
| medial_orbitofrontal_L | 50 | 5 | 14 | No |
| medial_orbitofrontal_R | 50 | 6 | 13 | No |
| middle_temporal_L | 50 | 11 | 12 | Yes |
| middle_temporal_R | 50 | 18 | 3 | No |
| parahippocampal_L | 50 | 11 | 13 | Yes |
| parahippocampal_R | 50 | 16 | 7 | No |
| paracentral_L | 50 | 13 | 9 | Yes |
| paracentral_R | 50 | 14 | 6 | No |
| pars_opercularis_L | 50 | 8 | 11 | Yes |
| pars_opercularis_R | 50 | 7 | 9 | No |
| pars_orbitalis_L | 48 | 5 | 7 | No |
| pars_orbitalis_R | 40 | 6 | 5 | No |
| pars_triangularis_L | 50 | 5 | 12 | No |
| pars_triangularis_R | 50 | 7 | 8 | No |
| pericalcarine_L | 22 | 4 | 0 | No |
| pericalcarine_R | 17 | 4 | 0 | No |
| postcentral_L | 50 | 10 | 8 | Yes |
| postcentral_R | 50 | 15 | 3 | No |
| posterior_cingulate_L | 50 | 7 | 14 | No |
| posterior_cingulate_R | 50 | 8 | 12 | Yes |
| precentral_L | 50 | 10 | 9 | Yes |
| precentral_R | 50 | 11 | 5 | No |
| precuneus_L | 50 | 18 | 7 | No |
| precuneus_R | 50 | 20 | 3 | No |

| | | | | |
|---|---|---|---|---|
| rostral_anterior_cingulate_L | 50 | 3 | 19 | No |
| rostral_anterior_cingulate_R | 50 | 4 | 16 | No |
| rostral_middle_frontal_L | 50 | 6 | 13 | No |
| rostral_middle_frontal_R | 50 | 8 | 10 | Yes |
| superior_frontal_L | 50 | 4 | 12 | No |
| superior_frontal_R | 50 | 8 | 8 | Yes |
| superior_parietal_L | 50 | 20 | 5 | No |
| superior_parietal_R | 50 | 25 | 2 | No |
| superior_temporal_L | 50 | 10 | 13 | Yes |
| superior_temporal_R | 50 | 11 | 6 | No |
| supramarginal_L | 50 | 12 | 8 | Yes |
| supramarginal_R | 50 | 17 | 5 | No |
| insula_L | 50 | 5 | 19 | No |
| insula_R | 50 | 4 | 7 | No |

**Caption:** Number of subjects contributing to the control, low-RT dose (≤15 Gy), and high-RT dose (≥40 Gy) groups for each ROI in the three-group RT dose comparison. Group sample sizes are reported as n and vary by ROI because RT dose grouping was defined on an ROI-specific basis and only subjects with usable ROI-wise CBF and RT dose data contributed. Eligibility for formal ANCOVA inference required at least 8 subjects per group. Gy, gray; ROI, region of interest.

#### Table S11. ROI-wise three-group ANCOVA of regional CBF by RT dose exposure group

| ROI | n control | n low RT dose | n high RT dose | Adjusted mean control | Adjusted mean low dose | Adjusted mean high dose | Contrast | Estimate | SE | t | p | q (FDR) |
|---|---|---|---|---|---|---|---|---|---|---|---|---|
| precentral_L | 50 | 10 | 9 | 115.5 | 112.1 | 88.2 | control vs high-RT dose | -27.2 | 5.6 | -4.84 | <0.001 | <0.001 |
| caudal_middle_frontal_L | 50 | 10 | 11 | 118.0 | 109.6 | 89.3 | control vs high-RT dose | -28.8 | 6.5 | -4.42 | <0.001 | 0.001 |
| rostral_middle_frontal_R | 50 | 8 | 10 | 109.7 | 98.6 | 90.7 | control vs high-RT dose | -19.1 | 5.5 | -3.49 | <0.001 | 0.016 |
| precentral_L | 50 | 10 | 9 | 115.5 | 112.1 | 88.2 | high-RT dose vs low-RT dose | 23.8 | 7.2 | 3.31 | 0.002 | 0.021 |
| postcentral_L | 50 | 10 | 8 | 104.0 | 100.7 | 86.8 | control vs high-RT dose | -17.3 | 5.8 | -2.98 | 0.004 | 0.044 |
| inferior_temporal_L | 50 | 15 | 9 | 71.7 | 60.5 | 66.7 | control vs low-RT dose | -11.2 | 3.9 | -2.88 | 0.005 | 0.048 |

| | | | | | | | | | | | | |
|---|---|---|---|---|---|---|---|---|---|---|---|---|
| superior_frontal_R | 50 | 8 | 8 | 103.5 | 91.2 | 87.5 | control vs high-RT dose | -16.0 | 5.8 | -2.75 | 0.008 | 0.061 |
| caudal_middle_frontal_R | 50 | 10 | 8 | 116.8 | 104.5 | 98.7 | control vs high-RT dose | -18.2 | 6.8 | -2.66 | 0.010 | 0.068 |
| caudal_middle_frontal_L | 50 | 10 | 11 | 118.0 | 109.6 | 89.3 | high-RT dose vs low-RT dose | 20.4 | 8.6 | 2.37 | 0.021 | 0.124 |
| hippocampus_R | 50 | 11 | 8 | 80.4 | 69.2 | 80.3 | control vs low-RT dose | -11.2 | 4.8 | -2.32 | 0.023 | 0.126 |
| superior_frontal_R | 50 | 8 | 8 | 103.5 | 91.2 | 87.5 | control vs low-RT dose | -12.3 | 5.9 | -2.09 | 0.041 | 0.202 |
| middle_temporal_L | 50 | 11 | 12 | 99.1 | 89.0 | 89.3 | control vs high-RT dose | -9.8 | 5.0 | -1.98 | 0.052 | 0.209 |
| middle_temporal_L | 50 | 11 | 12 | 99.1 | 89.0 | 89.3 | control vs low-RT dose | -10.1 | 5.1 | -1.98 | 0.052 | 0.209 |
| caudal_middle_frontal_R | 50 | 10 | 8 | 116.8 | 104.5 | 98.7 | control vs low-RT dose | -12.3 | 6.3 | -1.96 | 0.054 | 0.209 |
| postcentral_L | 50 | 10 | 8 | 104.0 | 100.7 | 86.8 | high-RT dose vs low-RT dose | 13.9 | 7.3 | 1.91 | 0.060 | 0.217 |
| rostral_middle_frontal_R | 50 | 8 | 10 | 109.7 | 98.6 | 90.7 | control vs low-RT dose | -11.1 | 6.0 | -1.84 | 0.070 | 0.231 |
| parahippocampal_L | 50 | 11 | 13 | 98.8 | 88.1 | 95.2 | control vs low-RT dose | -10.8 | 5.9 | -1.82 | 0.073 | 0.231 |
| fusiform_L | 50 | 16 | 8 | 94.8 | 87.8 | 93.5 | control vs low-RT dose | -7.0 | 4.1 | -1.71 | 0.092 | 0.276 |
| hippocampus_R | 50 | 11 | 8 | 80.4 | 69.2 | 80.3 | high-RT dose vs low-RT dose | -11.0 | 6.7 | -1.64 | 0.107 | 0.286 |
| entorhinal_L | 50 | 12 | 11 | 60.2 | 53.1 | 57.9 | control vs low-RT dose | -7.0 | 4.3 | -1.63 | 0.108 | 0.286 |
| superior_temporal_L | 50 | 10 | 13 | 96.3 | 90.7 | 88.9 | control vs high-RT dose | -7.3 | 4.5 | -1.61 | 0.111 | 0.286 |
| supramarginal_L | 50 | 12 | 8 | 111.7 | 106.9 | 103.8 | control vs high-RT dose | -7.9 | 5.9 | -1.35 | 0.183 | 0.447 |
| pars_opercularis_L | 50 | 8 | 11 | 116.3 | 123.6 | 109.0 | high-RT | 14.6 | 11.0 | 1.32 | 0.190 | 0.447 |

| | | | | | | | | | | | | |
|---|---|---|---|---|---|---|---|---|---|---|---|---|
| | | | | | | | dose vs low-RT dose | | | | | |
| caudal_middle_frontal_L | 50 | 10 | 11 | 118.0 | 109.6 | 89.3 | control vs low-RT dose | -8.4 | 6.8 | -1.24 | 0.221 | 0.498 |
| inferior_temporal_L | 50 | 15 | 9 | 71.7 | 60.5 | 66.7 | high-RT dose vs low-RT dose | -6.2 | 5.6 | -1.10 | 0.274 | 0.578 |
| superior_temporal_L | 50 | 10 | 13 | 96.3 | 90.7 | 88.9 | control vs low-RT dose | -5.6 | 5.1 | -1.09 | 0.278 | 0.578 |
| rostral_middle_frontal_R | 50 | 8 | 10 | 109.7 | 98.6 | 90.7 | high-RT dose vs low-RT dose | 8.0 | 7.5 | 1.06 | 0.293 | 0.586 |
| inferior_temporal_L | 50 | 15 | 9 | 71.7 | 60.5 | 66.7 | control vs high-RT dose | -5.0 | 4.8 | -1.03 | 0.306 | 0.586 |
| posterior_cingulate_R | 50 | 8 | 12 | 115.8 | 108.7 | 111.7 | control vs low-RT dose | -7.1 | 7.2 | -0.98 | 0.333 | 0.586 |
| parahippocampal_L | 50 | 11 | 13 | 98.8 | 88.1 | 95.2 | high-RT dose vs low-RT dose | -7.1 | 7.3 | -0.97 | 0.333 | 0.586 |
| supramarginal_L | 50 | 12 | 8 | 111.7 | 106.9 | 103.8 | control vs low-RT dose | -4.8 | 5.0 | -0.96 | 0.342 | 0.586 |
| pars_opercularis_L | 50 | 8 | 11 | 116.3 | 123.6 | 109.0 | control vs high-RT dose | -7.3 | 7.8 | -0.94 | 0.348 | 0.586 |
| fusiform_L | 50 | 16 | 8 | 94.8 | 87.8 | 93.5 | high-RT dose vs low-RT dose | -5.8 | 6.2 | -0.93 | 0.358 | 0.586 |
| entorhinal_L | 50 | 12 | 11 | 60.2 | 53.1 | 57.9 | high-RT dose vs low-RT dose | -4.7 | 5.6 | -0.84 | 0.402 | 0.638 |
| pars_opercularis_L | 50 | 8 | 11 | 116.3 | 123.6 | 109.0 | control vs low-RT dose | 7.2 | 9.0 | 0.80 | 0.427 | 0.658 |
| paracentral_L | 50 | 13 | 9 | 102.8 | 99.0 | 102.3 | control vs low-RT dose | -3.8 | 5.6 | -0.69 | 0.495 | 0.702 |
| caudal_middle_frontal_R | 50 | 10 | 8 | 116.8 | 104.5 | 98.7 | high-RT dose vs low-RT dose | 5.8 | 8.6 | 0.68 | 0.498 | 0.702 |
| posterior_cingulate_R | 50 | 8 | 12 | 115.8 | 108.7 | 111.7 | control vs high-RT dose | -4.1 | 6.1 | -0.67 | 0.505 | 0.702 |
| parahippocampal_L | 50 | 11 | 13 | 98.8 | 88.1 | 95.2 | control vs | -3.7 | 5.5 | -0.67 | 0.507 | 0.702 |

| | | | | | | | | | | | | |
|---|---|---|---|---|---|---|---|---|---|---|---|---|
| | | | | | | | high-RT dose | | | | | |
| precentral_L | 50 | 10 | 9 | 115.5 | 112.1 | 88.2 | control vs low-RT dose | -3.4 | 5.4 | -0.63 | 0.532 | 0.706 |
| postcentral_L | 50 | 10 | 8 | 104.0 | 100.7 | 86.8 | control vs low-RT dose | -3.3 | 5.3 | -0.62 | 0.536 | 0.706 |
| isthmus_cingulate_L | 50 | 8 | 8 | 122.0 | 118.3 | 118.3 | control vs high-RT dose | -3.7 | 6.7 | -0.54 | 0.588 | 0.740 |
| isthmus_cingulate_L | 50 | 8 | 8 | 122.0 | 118.3 | 118.3 | control vs low-RT dose | -3.7 | 6.8 | -0.54 | 0.590 | 0.740 |
| entorhinal_L | 50 | 12 | 11 | 60.2 | 53.1 | 57.9 | control vs high-RT dose | -2.3 | 4.5 | -0.52 | 0.608 | 0.746 |
| superior_frontal_R | 50 | 8 | 8 | 103.5 | 91.2 | 87.5 | high-RT dose vs low-RT dose | 3.7 | 7.7 | 0.48 | 0.634 | 0.761 |
| supramarginal_L | 50 | 12 | 8 | 111.7 | 106.9 | 103.8 | high-RT dose vs low-RT dose | 3.1 | 7.1 | 0.44 | 0.661 | 0.775 |
| paracentral_L | 50 | 13 | 9 | 102.8 | 99.0 | 102.3 | high-RT dose vs low-RT dose | -3.3 | 7.8 | -0.42 | 0.674 | 0.775 |
| posterior_cingulate_R | 50 | 8 | 12 | 115.8 | 108.7 | 111.7 | high-RT dose vs low-RT dose | -3.0 | 8.7 | -0.34 | 0.731 | 0.823 |
| superior_temporal_L | 50 | 10 | 13 | 96.3 | 90.7 | 88.9 | high-RT dose vs low-RT dose | 1.7 | 6.2 | 0.28 | 0.778 | 0.858 |
| fusiform_L | 50 | 16 | 8 | 94.8 | 87.8 | 93.5 | control vs high-RT dose | -1.2 | 5.5 | -0.23 | 0.821 | 0.887 |
| paracentral_L | 50 | 13 | 9 | 102.8 | 99.0 | 102.3 | control vs high-RT dose | -0.5 | 6.5 | -0.08 | 0.935 | 0.990 |
| middle_temporal_L | 50 | 11 | 12 | 99.1 | 89.0 | 89.3 | high-RT dose vs low-RT dose | -0.3 | 6.4 | -0.05 | 0.958 | 0.992 |
| hippocampus_R | 50 | 11 | 8 | 80.4 | 69.2 | 80.3 | control vs high-RT dose | -0.2 | 5.5 | -0.03 | 0.974 | 0.992 |
| isthmus_cingulate_L | 50 | 8 | 8 | 122.0 | 118.3 | 118.3 | high-RT dose vs | 0.0 | 8.9 | 0.00 | 0.997 | 0.997 |

| | | | | | | | |
|---|---|---|---|---|---|---|---|
| | | | | | | | low-RT dose |

**Caption:** ROI-wise three-group comparison of regional CBF by RT dose exposure group, reporting adjusted group means and pairwise contrasts from ANCOVA models (CBF ~ group + age + sex + DeepWM CBF). For each ROI, irradiated patients were classified into low-RT dose (≤15 Gy) or high-RT dose (≥40 Gy) groups on an ROI-specific basis and compared with healthy controls. The table includes the number of subjects per group (n), adjusted mean CBF values for each group, and pairwise contrast estimates with standard errors, t statistics, p-values, and Benjamini-Hochberg FDR-adjusted q-values. Only ROIs with ≥8 subjects per group were considered for formal inference. ANCOVA, analysis of covariance; CBF, cerebral blood flow; DeepWM, deep white matter; FDR, false discovery rate; ROI, region of interest; q, Benjamini-Hochberg adjusted p-value.

#### Table S12. Partial Spearman correlations between regional DKT perfusion and clinical variables in irradiated patients

| **Region** | **Predictor** | **n** | **Partial Spearman ρ** | **p** | **q (BH-FDR within predictor)** | **Significant q<0.05** |
|---|---|---|---|---|---|---|
| lateral_orbitofrontal_L | Chemotherapy (0/1) | 32 | -0.412 | 0.026 | 0.919 | No |
| precentral_L | Chemotherapy (0/1) | 33 | -0.317 | 0.088 | 0.919 | No |
| caudal_anterior_cingulate_L | Chemotherapy (0/1) | 28 | -0.324 | 0.114 | 0.919 | No |
| superior_parietal_L | Chemotherapy (0/1) | 33 | -0.249 | 0.184 | 0.919 | No |
| postcentral_L | Chemotherapy (0/1) | 33 | -0.243 | 0.196 | 0.919 | No |
| fusiform_L | Chemotherapy (0/1) | 33 | 0.233 | 0.216 | 0.919 | No |
| lateral_occipital_L | Chemotherapy (0/1) | 33 | -0.224 | 0.234 | 0.919 | No |
| insula_L | Chemotherapy (0/1) | 33 | -0.224 | 0.235 | 0.919 | No |
| caudal_middle_frontal_R | Chemotherapy (0/1) | 33 | -0.222 | 0.238 | 0.919 | No |
| thalamus_proper_R | Chemotherapy (0/1) | 33 | -0.219 | 0.245 | 0.919 | No |
| superior_frontal_R | Chemotherapy (0/1) | 33 | -0.215 | 0.254 | 0.919 | No |
| pars_triangularis_L | Chemotherapy (0/1) | 29 | -0.227 | 0.264 | 0.919 | No |
| posterior_cingulate_R | Chemotherapy (0/1) | 32 | -0.213 | 0.268 | 0.919 | No |
| paracentral_L | Chemotherapy (0/1) | 33 | -0.206 | 0.274 | 0.919 | No |
| entorhinal_L | Chemotherapy (0/1) | 30 | 0.210 | 0.292 | 0.919 | No |
| superior_parietal_R | Chemotherapy (0/1) | 33 | -0.181 | 0.338 | 0.919 | No |
| inferior_parietal_L | Chemotherapy (0/1) | 33 | -0.179 | 0.343 | 0.919 | No |
| putamen_L | Chemotherapy (0/1) | 31 | -0.183 | 0.352 | 0.919 | No |
| isthmus_cingulate_L | Chemotherapy (0/1) | 32 | -0.175 | 0.363 | 0.919 | No |
| rostral_middle_frontal_L | Chemotherapy (0/1) | 31 | -0.178 | 0.364 | 0.919 | No |
| caudate_L | Chemotherapy (0/1) | 29 | -0.183 | 0.372 | 0.919 | No |
| rostral_anterior_cingulate_L | Chemotherapy (0/1) | 31 | -0.173 | 0.378 | 0.919 | No |

| | | | | | | |
|---|---|---|---|---|---|---|
| rostral_middle_frontal_R | Chemotherapy (0/1) | 33 | -0.165 | 0.382 | 0.919 | No |
| precuneus_L | Chemotherapy (0/1) | 33 | -0.165 | 0.383 | 0.919 | No |
| entorhinal_R | Chemotherapy (0/1) | 30 | 0.169 | 0.399 | 0.919 | No |
| caudal_middle_frontal_L | Chemotherapy (0/1) | 31 | -0.165 | 0.401 | 0.919 | No |
| superior_frontal_L | Chemotherapy (0/1) | 33 | -0.159 | 0.403 | 0.919 | No |
| lateral_orbitofrontal_R | Chemotherapy (0/1) | 33 | -0.148 | 0.435 | 0.919 | No |
| putamen_R | Chemotherapy (0/1) | 32 | -0.151 | 0.435 | 0.919 | No |
| inferior_temporal_R | Chemotherapy (0/1) | 33 | 0.147 | 0.437 | 0.919 | No |
| caudate_R | Chemotherapy (0/1) | 31 | -0.150 | 0.445 | 0.919 | No |
| hippocampus_L | Chemotherapy (0/1) | 28 | 0.156 | 0.455 | 0.919 | No |
| hippocampus_R | Chemotherapy (0/1) | 32 | -0.140 | 0.468 | 0.919 | No |
| superior_temporal_R | Chemotherapy (0/1) | 33 | -0.136 | 0.473 | 0.919 | No |
| parahippocampal_L | Chemotherapy (0/1) | 32 | 0.129 | 0.504 | 0.942 | No |
| postcentral_R | Chemotherapy (0/1) | 33 | -0.119 | 0.532 | 0.942 | No |
| supramarginal_L | Chemotherapy (0/1) | 32 | -0.115 | 0.554 | 0.942 | No |
| lateral_occipital_R | Chemotherapy (0/1) | 33 | 0.105 | 0.583 | 0.942 | No |
| pars_opercularis_R | Chemotherapy (0/1) | 31 | 0.108 | 0.585 | 0.942 | No |
| pars_orbitalis_R | Chemotherapy (0/1) | 24 | -0.121 | 0.600 | 0.942 | No |
| middle_temporal_L | Chemotherapy (0/1) | 33 | -0.095 | 0.616 | 0.942 | No |
| pars_orbitalis_L | Chemotherapy (0/1) | 21 | -0.123 | 0.627 | 0.942 | No |
| superior_temporal_L | Chemotherapy (0/1) | 33 | -0.080 | 0.673 | 0.942 | No |
| isthmus_cingulate_R | Chemotherapy (0/1) | 32 | 0.081 | 0.676 | 0.942 | No |
| thalamus_proper_L | Chemotherapy (0/1) | 33 | -0.072 | 0.705 | 0.942 | No |
| inferior_parietal_R | Chemotherapy (0/1) | 33 | -0.071 | 0.709 | 0.942 | No |
| paracentral_R | Chemotherapy (0/1) | 33 | -0.069 | 0.716 | 0.942 | No |
| medial_orbitofrontal_R | Chemotherapy (0/1) | 32 | -0.070 | 0.719 | 0.942 | No |
| caudal_anterior_cingulate_R | Chemotherapy (0/1) | 29 | -0.073 | 0.723 | 0.942 | No |
| middle_temporal_R | Chemotherapy (0/1) | 33 | -0.066 | 0.731 | 0.942 | No |
| lingual_L | Chemotherapy (0/1) | 33 | -0.064 | 0.738 | 0.942 | No |
| lingual_R | Chemotherapy (0/1) | 33 | 0.061 | 0.750 | 0.942 | No |
| pars_triangularis_R | Chemotherapy (0/1) | 31 | 0.060 | 0.761 | 0.942 | No |
| posterior_cingulate_L | Chemotherapy (0/1) | 33 | -0.056 | 0.770 | 0.942 | No |
| cuneus_R | Chemotherapy (0/1) | 33 | 0.046 | 0.809 | 0.955 | No |
| medial_orbitofrontal_L | Chemotherapy (0/1) | 32 | 0.041 | 0.833 | 0.955 | No |
| parahippocampal_R | Chemotherapy (0/1) | 33 | 0.040 | 0.834 | 0.955 | No |
| pars_opercularis_L | Chemotherapy (0/1) | 26 | -0.044 | 0.841 | 0.955 | No |

| | | | | | | |
|---|---|---|---|---|---|---|
| precentral_R | Chemotherapy (0/1) | 33 | -0.031 | 0.870 | 0.955 | No |
| inferior_temporal_L | Chemotherapy (0/1) | 33 | 0.031 | 0.871 | 0.955 | No |
| supramarginal_R | Chemotherapy (0/1) | 33 | -0.026 | 0.890 | 0.955 | No |
| cuneus_L | Chemotherapy (0/1) | 28 | 0.027 | 0.897 | 0.955 | No |
| insula_R | Chemotherapy (0/1) | 31 | 0.019 | 0.922 | 0.966 | No |
| fusiform_R | Chemotherapy (0/1) | 33 | 0.010 | 0.956 | 0.982 | No |
| precuneus_R | Chemotherapy (0/1) | 33 | -0.008 | 0.967 | 0.982 | No |
| rostral_anterior_cingulate_R | Chemotherapy (0/1) | 29 | 0.003 | 0.988 | 0.988 | No |
| lateral_orbitofrontal_L | Time since RT (years) | 32 | 0.340 | 0.071 | 0.988 | No |
| lingual_R | Time since RT (years) | 33 | -0.309 | 0.097 | 0.988 | No |
| lingual_L | Time since RT (years) | 33 | -0.249 | 0.185 | 0.988 | No |
| pars_triangularis_R | Time since RT (years) | 31 | 0.253 | 0.194 | 0.988 | No |
| hippocampus_L | Time since RT (years) | 28 | -0.261 | 0.207 | 0.988 | No |
| putamen_R | Time since RT (years) | 32 | 0.226 | 0.239 | 0.988 | No |
| parahippocampal_R | Time since RT (years) | 33 | -0.217 | 0.249 | 0.988 | No |
| isthmus_cingulate_L | Time since RT (years) | 32 | 0.212 | 0.269 | 0.988 | No |
| cuneus_L | Time since RT (years) | 28 | -0.228 | 0.273 | 0.988 | No |
| parahippocampal_L | Time since RT (years) | 32 | -0.204 | 0.288 | 0.988 | No |
| insula_R | Time since RT (years) | 31 | 0.199 | 0.310 | 0.988 | No |
| paracentral_L | Time since RT (years) | 33 | -0.176 | 0.352 | 0.988 | No |
| inferior_parietal_L | Time since RT (years) | 33 | 0.172 | 0.363 | 0.988 | No |
| postcentral_L | Time since RT (years) | 33 | -0.162 | 0.392 | 0.988 | No |
| superior_temporal_R | Time since RT (years) | 33 | 0.160 | 0.400 | 0.988 | No |
| pars_orbitalis_L | Time since RT (years) | 21 | 0.209 | 0.406 | 0.988 | No |
| lateral_occipital_L | Time since RT (years) | 33 | 0.145 | 0.444 | 0.988 | No |
| rostral_anterior_cingulate_R | Time since RT (years) | 29 | 0.154 | 0.454 | 0.988 | No |
| caudate_L | Time since RT (years) | 29 | 0.154 | 0.454 | 0.988 | No |
| entorhinal_L | Time since RT (years) | 30 | -0.145 | 0.470 | 0.988 | No |
| precentral_L | Time since RT (years) | 33 | -0.132 | 0.486 | 0.988 | No |
| inferior_parietal_R | Time since RT (years) | 33 | 0.129 | 0.495 | 0.988 | No |
| pars_opercularis_R | Time since RT (years) | 31 | 0.132 | 0.505 | 0.988 | No |
| cuneus_R | Time since RT (years) | 33 | -0.124 | 0.513 | 0.988 | No |
| lateral_orbitofrontal_R | Time since RT (years) | 33 | -0.122 | 0.522 | 0.988 | No |
| superior_temporal_L | Time since RT (years) | 33 | -0.103 | 0.589 | 0.988 | No |
| paracentral_R | Time since RT (years) | 33 | -0.099 | 0.602 | 0.988 | No |
| posterior_cingulate_R | Time since RT (years) | 32 | 0.094 | 0.627 | 0.988 | No |

| | | | | | | |
|---|---|---|---|---|---|---|
| caudal_middle_frontal_R | Time since RT (years) | 33 | -0.088 | 0.645 | 0.988 | No |
| caudate_R | Time since RT (years) | 31 | 0.091 | 0.646 | 0.988 | No |
| insula_L | Time since RT (years) | 33 | 0.085 | 0.655 | 0.988 | No |
| fusiform_L | Time since RT (years) | 33 | -0.085 | 0.657 | 0.988 | No |
| superior_frontal_L | Time since RT (years) | 33 | 0.080 | 0.673 | 0.988 | No |
| superior_parietal_R | Time since RT (years) | 33 | -0.078 | 0.684 | 0.988 | No |
| thalamus_proper_R | Time since RT (years) | 33 | 0.078 | 0.684 | 0.988 | No |
| superior_parietal_L | Time since RT (years) | 33 | 0.071 | 0.709 | 0.988 | No |
| inferior_temporal_R | Time since RT (years) | 33 | -0.070 | 0.712 | 0.988 | No |
| thalamus_proper_L | Time since RT (years) | 33 | 0.065 | 0.734 | 0.988 | No |
| superior_frontal_R | Time since RT (years) | 33 | 0.064 | 0.738 | 0.988 | No |
| pars_opercularis_L | Time since RT (years) | 26 | -0.073 | 0.741 | 0.988 | No |
| pars_triangularis_L | Time since RT (years) | 29 | 0.065 | 0.753 | 0.988 | No |
| rostral_middle_frontal_L | Time since RT (years) | 31 | 0.062 | 0.755 | 0.988 | No |
| isthmus_cingulate_R | Time since RT (years) | 32 | 0.060 | 0.759 | 0.988 | No |
| pars_orbitalis_R | Time since RT (years) | 24 | -0.070 | 0.764 | 0.988 | No |
| posterior_cingulate_L | Time since RT (years) | 33 | 0.056 | 0.770 | 0.988 | No |
| caudal_anterior_cingulate_R | Time since RT (years) | 29 | 0.058 | 0.777 | 0.988 | No |
| precentral_R | Time since RT (years) | 33 | -0.049 | 0.798 | 0.988 | No |
| postcentral_R | Time since RT (years) | 33 | -0.046 | 0.809 | 0.988 | No |
| precuneus_L | Time since RT (years) | 33 | 0.043 | 0.823 | 0.988 | No |
| caudal_anterior_cingulate_L | Time since RT (years) | 28 | -0.043 | 0.837 | 0.988 | No |
| supramarginal_R | Time since RT (years) | 33 | 0.036 | 0.851 | 0.988 | No |
| putamen_L | Time since RT (years) | 31 | 0.037 | 0.852 | 0.988 | No |
| rostral_anterior_cingulate_L | Time since RT (years) | 31 | 0.036 | 0.857 | 0.988 | No |
| rostral_middle_frontal_R | Time since RT (years) | 33 | 0.032 | 0.868 | 0.988 | No |
| middle_temporal_L | Time since RT (years) | 33 | -0.022 | 0.908 | 0.988 | No |
| medial_orbitofrontal_L | Time since RT (years) | 32 | -0.020 | 0.917 | 0.988 | No |
| fusiform_R | Time since RT (years) | 33 | -0.017 | 0.929 | 0.988 | No |
| medial_orbitofrontal_R | Time since RT (years) | 32 | 0.017 | 0.931 | 0.988 | No |
| entorhinal_R | Time since RT (years) | 30 | 0.017 | 0.932 | 0.988 | No |
| middle_temporal_R | Time since RT (years) | 33 | 0.016 | 0.935 | 0.988 | No |
| precuneus_R | Time since RT (years) | 33 | 0.014 | 0.943 | 0.988 | No |
| inferior_temporal_L | Time since RT (years) | 33 | 0.010 | 0.957 | 0.988 | No |
| lateral_occipital_R | Time since RT (years) | 33 | -0.010 | 0.959 | 0.988 | No |
| supramarginal_L | Time since RT (years) | 32 | -0.008 | 0.969 | 0.988 | No |

| | | | | | | |
|---|---|---|---|---|---|---|
| caudal_middle_frontal_L | Time since RT (years) | 31 | 0.007 | 0.973 | 0.988 | No |
| hippocampus_R | Time since RT (years) | 32 | 0.003 | 0.990 | 0.990 | No |
| parahippocampal_R | Tumor volume ($cm^3$) | 33 | 0.387 | 0.034 | 0.987 | No |
| pars_triangularis_R | Tumor volume ($cm^3$) | 31 | -0.380 | 0.046 | 0.987 | No |
| superior_parietal_R | Tumor volume ($cm^3$) | 33 | -0.353 | 0.056 | 0.987 | No |
| hippocampus_L | Tumor volume ($cm^3$) | 28 | 0.357 | 0.080 | 0.987 | No |
| lingual_L | Tumor volume ($cm^3$) | 33 | 0.263 | 0.160 | 0.987 | No |
| pars_opercularis_L | Tumor volume ($cm^3$) | 26 | 0.268 | 0.216 | 0.987 | No |
| thalamus_proper_R | Tumor volume ($cm^3$) | 33 | 0.225 | 0.233 | 0.987 | No |
| cuneus_L | Tumor volume ($cm^3$) | 28 | 0.246 | 0.236 | 0.987 | No |
| superior_temporal_L | Tumor volume ($cm^3$) | 33 | 0.220 | 0.243 | 0.987 | No |
| fusiform_R | Tumor volume ($cm^3$) | 33 | 0.218 | 0.247 | 0.987 | No |
| lateral_occipital_R | Tumor volume ($cm^3$) | 33 | -0.200 | 0.289 | 0.987 | No |
| parahippocampal_L | Tumor volume ($cm^3$) | 32 | 0.199 | 0.300 | 0.987 | No |
| entorhinal_R | Tumor volume ($cm^3$) | 30 | -0.200 | 0.318 | 0.987 | No |
| paracentral_R | Tumor volume ($cm^3$) | 33 | 0.184 | 0.330 | 0.987 | No |
| postcentral_R | Tumor volume ($cm^3$) | 33 | -0.172 | 0.363 | 0.987 | No |
| putamen_R | Tumor volume ($cm^3$) | 32 | -0.159 | 0.409 | 0.987 | No |
| isthmus_cingulate_R | Tumor volume ($cm^3$) | 32 | 0.152 | 0.432 | 0.987 | No |
| rostral_middle_frontal_R | Tumor volume ($cm^3$) | 33 | -0.148 | 0.434 | 0.987 | No |
| paracentral_L | Tumor volume ($cm^3$) | 33 | 0.144 | 0.447 | 0.987 | No |
| inferior_temporal_R | Tumor volume ($cm^3$) | 33 | 0.142 | 0.454 | 0.987 | No |
| fusiform_L | Tumor volume ($cm^3$) | 33 | 0.138 | 0.467 | 0.987 | No |
| supramarginal_R | Tumor volume ($cm^3$) | 33 | 0.126 | 0.507 | 0.987 | No |
| pars_triangularis_L | Tumor volume ($cm^3$) | 29 | -0.136 | 0.508 | 0.987 | No |
| rostral_anterior_cingulate_R | Tumor volume ($cm^3$) | 29 | -0.134 | 0.513 | 0.987 | No |
| lingual_R | Tumor volume ($cm^3$) | 33 | 0.119 | 0.532 | 0.987 | No |
| lateral_orbitofrontal_L | Tumor volume ($cm^3$) | 32 | -0.119 | 0.540 | 0.987 | No |
| inferior_parietal_L | Tumor volume ($cm^3$) | 33 | 0.115 | 0.543 | 0.987 | No |
| superior_frontal_L | Tumor volume ($cm^3$) | 33 | 0.109 | 0.565 | 0.987 | No |
| isthmus_cingulate_L | Tumor volume ($cm^3$) | 32 | -0.110 | 0.570 | 0.987 | No |
| entorhinal_L | Tumor volume ($cm^3$) | 30 | 0.107 | 0.594 | 0.987 | No |
| lateral_occipital_L | Tumor volume ($cm^3$) | 33 | 0.100 | 0.600 | 0.987 | No |
| caudal_middle_frontal_L | Tumor volume ($cm^3$) | 31 | -0.098 | 0.621 | 0.987 | No |
| middle_temporal_L | Tumor volume ($cm^3$) | 33 | 0.089 | 0.641 | 0.987 | No |
| caudal_anterior_cingulate_L | Tumor volume ($cm^3$) | 28 | -0.085 | 0.687 | 0.987 | No |

| | | | | | | |
|---|---|---|---|---|---|---|
| posterior_cingulate_L | Tumor volume ($cm^3$) | 33 | 0.073 | 0.701 | 0.987 | No |
| precentral_R | Tumor volume ($cm^3$) | 33 | -0.071 | 0.711 | 0.987 | No |
| precentral_L | Tumor volume ($cm^3$) | 33 | -0.070 | 0.712 | 0.987 | No |
| inferior_parietal_R | Tumor volume ($cm^3$) | 33 | -0.067 | 0.724 | 0.987 | No |
| supramarginal_L | Tumor volume ($cm^3$) | 32 | 0.068 | 0.727 | 0.987 | No |
| putamen_L | Tumor volume ($cm^3$) | 31 | 0.068 | 0.732 | 0.987 | No |
| caudate_L | Tumor volume ($cm^3$) | 29 | -0.067 | 0.745 | 0.987 | No |
| pars_orbitalis_R | Tumor volume ($cm^3$) | 24 | -0.075 | 0.745 | 0.987 | No |
| caudal_anterior_cingulate_R | Tumor volume ($cm^3$) | 29 | 0.067 | 0.746 | 0.987 | No |
| superior_temporal_R | Tumor volume ($cm^3$) | 33 | 0.057 | 0.764 | 0.987 | No |
| inferior_temporal_L | Tumor volume ($cm^3$) | 33 | 0.055 | 0.774 | 0.987 | No |
| caudal_middle_frontal_R | Tumor volume ($cm^3$) | 33 | -0.050 | 0.794 | 0.987 | No |
| medial_orbitofrontal_L | Tumor volume ($cm^3$) | 32 | 0.050 | 0.795 | 0.987 | No |
| rostral_anterior_cingulate_L | Tumor volume ($cm^3$) | 31 | 0.051 | 0.795 | 0.987 | No |
| pars_orbitalis_L | Tumor volume ($cm^3$) | 21 | -0.065 | 0.797 | 0.987 | No |
| lateral_orbitofrontal_R | Tumor volume ($cm^3$) | 33 | -0.048 | 0.802 | 0.987 | No |
| posterior_cingulate_R | Tumor volume ($cm^3$) | 32 | 0.046 | 0.814 | 0.987 | No |
| superior_frontal_R | Tumor volume ($cm^3$) | 33 | -0.043 | 0.820 | 0.987 | No |
| precuneus_R | Tumor volume ($cm^3$) | 33 | -0.040 | 0.836 | 0.987 | No |
| rostral_middle_frontal_L | Tumor volume ($cm^3$) | 31 | 0.040 | 0.839 | 0.987 | No |
| insula_R | Tumor volume ($cm^3$) | 31 | -0.039 | 0.846 | 0.987 | No |
| caudate_R | Tumor volume ($cm^3$) | 31 | 0.034 | 0.863 | 0.987 | No |
| pars_opercularis_R | Tumor volume ($cm^3$) | 31 | -0.028 | 0.886 | 0.987 | No |
| thalamus_proper_L | Tumor volume ($cm^3$) | 33 | 0.025 | 0.897 | 0.987 | No |
| superior_parietal_L | Tumor volume ($cm^3$) | 33 | -0.024 | 0.899 | 0.987 | No |
| middle_temporal_R | Tumor volume ($cm^3$) | 33 | 0.022 | 0.909 | 0.987 | No |
| postcentral_L | Tumor volume ($cm^3$) | 33 | 0.021 | 0.912 | 0.987 | No |
| precuneus_L | Tumor volume ($cm^3$) | 33 | -0.016 | 0.935 | 0.995 | No |
| insula_L | Tumor volume ($cm^3$) | 33 | 0.007 | 0.973 | 0.995 | No |
| hippocampus_R | Tumor volume ($cm^3$) | 32 | -0.004 | 0.982 | 0.995 | No |
| cuneus_R | Tumor volume ($cm^3$) | 33 | 0.001 | 0.995 | 0.995 | No |
| medial_orbitofrontal_R | Tumor volume ($cm^3$) | 32 | -0.001 | 0.995 | 0.995 | No |

**Caption**: ROI-wise partial Spearman correlations between unilateral DKT regional cerebral blood flow (CBF) and clinical variables (time since RT (years), tumor volume ($cm^3$), and chemotherapy exposure) in irradiated patients, adjusted for Age, sex, and DeepWM CBF. Only ROIs with at least 10 available observations were included. N denotes the number of irradiated patients contributing to each region-predictor analysis and may vary across rows because of region-specific exclusions or missing clinical data. Raw p-values and Benjamini-Hochberg false discovery rate (FDR)-adjusted q-values are reported within predictor across ROIs. Chemotherapy was coded as 0 (no) or 1 (yes). DKT, Desikan-Killiany-Tourville; CBF, cerebral blood flow; DeepWM, deep white matter; FDR, false discovery rate; q, Benjamini-Hochberg adjusted p-value.

**Table S13. A priori ROI cognition-CBF partial correlations in irradiated patients**

| Domain | ROI (DKT) | r | p | n | q |
|---|---|---|---|---|---|
| Attention/Processing Speed | caudal_middle_frontal_L | 0.439 | 0.013 | 31 | 0.162 |
| Attention/Processing Speed | caudal_middle_frontal_R | 0.180 | 0.317 | 33 | 0.447 |
| Attention/Processing Speed | inferior_parietal_L | 0.276 | 0.120 | 33 | 0.334 |
| Attention/Processing Speed | inferior_parietal_R | 0.127 | 0.481 | 33 | 0.503 |
| Attention/Processing Speed | lateral_occipital_L | 0.121 | 0.503 | 33 | 0.503 |
| Attention/Processing Speed | lateral_occipital_R | 0.203 | 0.258 | 33 | 0.442 |
| Attention/Processing Speed | precuneus_L | 0.288 | 0.104 | 33 | 0.334 |
| Attention/Processing Speed | precuneus_R | 0.123 | 0.494 | 33 | 0.503 |
| Attention/Processing Speed | rostral_middle_frontal_L | 0.249 | 0.177 | 31 | 0.354 |
| Attention/Processing Speed | rostral_middle_frontal_R | 0.173 | 0.335 | 33 | 0.447 |
| Attention/Processing Speed | superior_parietal_L | 0.274 | 0.123 | 33 | 0.334 |
| Attention/Processing Speed | superior_parietal_R | 0.263 | 0.139 | 33 | 0.334 |
| Executive Function | caudal_anterior_cingulate_L | 0.402 | 0.034 | 28 | 0.271 |
| Executive Function | caudal_anterior_cingulate_R | -0.062 | 0.748 | 29 | 0.855 |
| Executive Function | caudal_middle_frontal_L | 0.412 | 0.021 | 31 | 0.271 |
| Executive Function | caudal_middle_frontal_R | 0.086 | 0.633 | 33 | 0.816 |
| Executive Function | insula_L | 0.286 | 0.106 | 33 | 0.339 |
| Executive Function | insula_R | 0.160 | 0.391 | 31 | 0.631 |
| Executive Function | lateral_orbitofrontal_L | 0.336 | 0.060 | 32 | 0.318 |
| Executive Function | lateral_orbitofrontal_R | 0.207 | 0.247 | 33 | 0.564 |
| Executive Function | medial_orbitofrontal_L | 0.143 | 0.434 | 32 | 0.631 |
| Executive Function | medial_orbitofrontal_R | 0.000 | 0.999 | 32 | 0.999 |
| Executive Function | rostral_anterior_cingulate_L | 0.256 | 0.165 | 31 | 0.440 |
| Executive Function | rostral_anterior_cingulate_R | 0.158 | 0.413 | 29 | 0.631 |
| Executive Function | rostral_middle_frontal_L | 0.300 | 0.101 | 31 | 0.339 |
| Executive Function | rostral_middle_frontal_R | 0.079 | 0.663 | 33 | 0.816 |
| Executive Function | thalamus_proper_L | 0.148 | 0.411 | 33 | 0.631 |
| Executive Function | thalamus_proper_R | -0.014 | 0.938 | 33 | 0.999 |
| Proxy IQ | caudal_anterior_cingulate_L | 0.356 | 0.063 | 28 | 0.188 |
| Proxy IQ | caudal_anterior_cingulate_R | 0.258 | 0.176 | 29 | 0.265 |

| | | | | | |
|---|---|---|---|---|---|
| Proxy IQ | caudal_middle_frontal_L | 0.435 | 0.014 | 31 | 0.058 |
| Proxy IQ | caudal_middle_frontal_R | 0.436 | 0.011 | 33 | 0.058 |
| Proxy IQ | insula_L | 0.123 | 0.494 | 33 | 0.494 |
| Proxy IQ | insula_R | 0.319 | 0.080 | 31 | 0.193 |
| Proxy IQ | rostral_anterior_cingulate_L | 0.204 | 0.272 | 31 | 0.326 |
| Proxy IQ | rostral_anterior_cingulate_R | 0.277 | 0.145 | 29 | 0.249 |
| Proxy IQ | rostral_middle_frontal_L | 0.213 | 0.249 | 31 | 0.326 |
| Proxy IQ | rostral_middle_frontal_R | 0.441 | 0.010 | 33 | 0.058 |
| Proxy IQ | thalamus_proper_L | 0.282 | 0.111 | 33 | 0.223 |
| Proxy IQ | thalamus_proper_R | 0.184 | 0.305 | 33 | 0.332 |
| Language | inferior_temporal_L | 0.176 | 0.327 | 33 | 0.509 |
| Language | inferior_temporal_R | 0.178 | 0.323 | 33 | 0.509 |
| Language | middle_temporal_L | 0.331 | 0.060 | 33 | 0.211 |
| Language | middle_temporal_R | 0.100 | 0.580 | 33 | 0.677 |
| Language | pars_opercularis_L | 0.315 | 0.117 | 26 | 0.235 |
| Language | pars_opercularis_R | -0.040 | 0.829 | 31 | 0.840 |
| Language | pars_orbitalis_L | 0.134 | 0.561 | 21 | 0.677 |
| Language | pars_orbitalis_R | -0.329 | 0.116 | 24 | 0.235 |
| Language | pars_triangularis_L | 0.361 | 0.054 | 29 | 0.211 |
| Language | pars_triangularis_R | -0.038 | 0.840 | 31 | 0.840 |
| Language | superior_temporal_L | 0.400 | 0.021 | 33 | 0.211 |
| Language | superior_temporal_R | 0.279 | 0.115 | 33 | 0.235 |
| Language | supramarginal_L | 0.353 | 0.048 | 32 | 0.211 |
| Language | supramarginal_R | 0.127 | 0.481 | 33 | 0.674 |
| Memory | entorhinal_L | 0.044 | 0.818 | 30 | 0.821 |
| Memory | entorhinal_R | 0.158 | 0.404 | 30 | 0.821 |
| Memory | hippocampus_L | -0.078 | 0.694 | 28 | 0.821 |
| Memory | hippocampus_R | 0.163 | 0.371 | 32 | 0.821 |
| Memory | isthmus_cingulate_L | 0.281 | 0.119 | 32 | 0.821 |
| Memory | isthmus_cingulate_R | -0.042 | 0.821 | 32 | 0.821 |
| Memory | medial_orbitofrontal_L | 0.043 | 0.815 | 32 | 0.821 |
| Memory | medial_orbitofrontal_R | -0.052 | 0.778 | 32 | 0.821 |
| Memory | parahippocampal_L | -0.096 | 0.601 | 32 | 0.821 |
| Memory | parahippocampal_R | -0.146 | 0.418 | 33 | 0.821 |
| Memory | posterior_cingulate_L | 0.275 | 0.122 | 33 | 0.821 |
| Memory | posterior_cingulate_R | 0.216 | 0.235 | 32 | 0.821 |
| Memory | thalamus_proper_L | 0.186 | 0.301 | 33 | 0.821 |
| Memory | thalamus_proper_R | 0.055 | 0.762 | 33 | 0.821 |
| Motor | paracentral_L | 0.253 | 0.155 | 33 | 0.186 |
| Motor | paracentral_R | 0.090 | 0.619 | 33 | 0.619 |
| Motor | postcentral_L | 0.653 | <0.001 | 33 | <0.001 |
| Motor | postcentral_R | 0.314 | 0.076 | 33 | 0.114 |
| Motor | precentral_L | 0.689 | <0.001 | 33 | <0.001 |
| Motor | precentral_R | 0.367 | 0.035 | 33 | 0.071 |

**Caption**: Partial Pearson correlations between ROI-wise CBF and cognitive domain w-scores were performed within the irradiated subgroup and adjusted for DeepWM CBF. Analyses were restricted to a priori domain-specific ROI sets. Benjamini-Hochberg FDR correction was applied within each cognitive domain. Exact p-values and within-domain Benjamini-Hochberg false discovery rate (FDR)-adjusted q-values are reported. N denotes the number of irradiated patients contributing to each ROI-domain analysis and may vary across rows because of ROI-specific exclusions or missing cognitive data. CBF, cerebral blood flow; DeepWM, deep white matter; DKT, Desikan-Killiany-Tourville atlas; FDR, false discovery rate; ROI, region of interest.
